# Automated Detection of Cerebral Microbleeds on MR images using Knowledge Distillation Framework

**DOI:** 10.1101/2021.11.15.21266376

**Authors:** Vaanathi Sundaresan, Christoph Arthofer, Giovanna Zamboni, Andrew G. Murchison, Robert A. Dineen, Peter M. Rothwell, Dorothee P. Auer, Chaoyue Wang, Karla L. Miller, Benjamin C. Tendler, Fidel Alfaro-Almagro, Stamatios N. Sotiropoulos, Nikola Sprigg, Ludovica Griffanti, Mark Jenkinson

## Abstract

Cerebral microbleeds (CMBs) are associated with white matter damage, various neurodegenerative and cerebrovascular diseases. CMBs occur as small, circular hypointense lesions on T2*-weighted gradient recalled echo (GRE) and susceptibility weighted imaging (SWI) images, and hyperintense on quantitative susceptibility mapping (QSM) images due to their paramagnetic nature. Accurate detection of CMBs would help to determine the CMB lesion count and distribution, which would be further useful to understand the clinical impact of CMBs and to obtain quantitative imaging biomarkers. In this work, we propose a fully automated, deep learning-based, 2-step algorithm, using structural and anatomical properties of CMBs from any single input image modality (e.g. GRE / SWI / QSM) for their accurate detection. Our method consists of an initial candidate detection step, that detects CMBs with high sensitivity and a candidate discrimination step using a knowledge distillation framework to classify CMB and non-CMB instances, followed by a morphological clean-up step. We used 4 datasets consisting of different modalities specified above, acquired using various protocols and with a variety of pathological and demographic characteristics. On cross-validation within datasets, our method achieved a cluster-wise true positive rate (TPR) over 90% with an average of less than 2 false positives per subject. Our method is flexible in terms of the input modality and provides comparable cluster-wise TPR and a better cluster-wise precision compared to existing state-of-the-art methods. When evaluated across different datasets, our method showed good generalisability with a cluster-wise TPR greater than 80% with different modalities.

## 1. Introduction

Cerebral microbleeds (CMBs) are haemosiderin deposits due to micro-haemorrhages in the brain. CMBs are found in subjects with cerebrovascular diseases, cognitive impairment and dementia, and also found in healthy elderly subjects. CMBs have been associated with white matter damage, various neurogenerative diseases including Alzheimer’s disease and cerebral amyloid angiopathy (CAA). The presence of CMBs has also been shown to increase the risk of symptomatic intracerebral haemorrhage (ICH) and stroke (Cordonnier et al., 2007). Identification of CMBs and determining their distribution could help in obtaining important biomarkers for various diseases (e.g. lobar CMBs and deep/infratentorial CMBs might indicate CAA and hypertensive vasculopathy respectively (Greenberg et al., 2009)). CMBs appear as small, circular, well defined hypointense lesions ranging from 2 to 10 mm in size on T2*-weighted gradient recalled echo (GRE) images. Due to the paramagnetic susceptibility properties of the iron content in the CMBs, modalities such as susceptibility weighted imaging (SWI) (Haacke et al., 2004) and quantitative susceptibility mapping (QSM) images (Liu et al., 2015) are useful in the identification of CMBs. While all the above modalities are derived from the same scan, they use different aspects of data - T2*-weighted GRE derived from magnitude only, QSM from phase only, and SWI derived from a combination of phase and magnitude. When compared to T2*-weighted GRE (T2*-GRE) images, CMBs appear more prominently on SWI images due to the blooming effect (Greenberg et al., 2009; Charidimou and Werring, 2011). Unlike T2*-GRE and SWI modalities, CMBs appear hyperintense on QSM images. Automated detection of CMBs is highly challenging due to their small size, contrast variations, sparse distribution and the presence of imaging artefacts (e.g. ringing effect, susceptibility artefacts at tissue interfaces). Additionally, the presence of various ‘CMB-like’ structures (or *mimics*) with diamagnetic (e.g. calcifications) and paramagnetic (e.g. micrometastases and haemorrhages) properties makes the accurate detection of CMBs very difficult (for the list of mimics and their description, refer to Greenberg et al. (2009)). While the use of SWI images generally improves the CMB contrast when compared to GRE magnitude images (Nandigam et al., 2009; Shams et al., 2015), SWI also enhances mimics with magnetic susceptibility differences (both diamagnetic and paramagnetic), making it difficult to identify true CMBs (Greenberg et al., 2009). QSM could be useful to accurately identify true CMBs since it allows to separate diamagnetic tissues (with negative susceptibility, appearing hypointense) from paramagnetic tissues (with positive susceptibility, appering hyperintense). On QSM images, CMBs appear hyperintense (unlike on other considered modalities) while diamagnetic mimics (e.g. calcifications) will appear hypointense (Rashid et al., 2021).

### 1.1. Existing literature

#### CMB detection

Various semi-automated and automated methods have been proposed for CMB detection. Most of the methods follow a common pattern with two steps: CMB candidate detection and post-processing to remove false positives (FPs). The first step generally achieves high sensitivity, while the second step is more challenging and leads to improvement in the precision. In the semi-automated methods, manual intervention has often been used in the cleaning-up step to remove FPs (Seghier et al., 2011; Barnes et al., 2011; van den Heuvel et al., 2016; Morrison et al., 2018). Occasionally, candidate detection (De Bresser et al., 2013; Lu et al., 2021a) and ground truth verification (Kuijf et al., 2012, 2013) also involves manual intervention. Manual detection of CMB candidates are extremely labour intensive, especially when done on large number of subjects (e.g., around 8000 subjects from the UK Biobank (Lu et al., 2021a)), and might increase the risk of observer error, given the large number of scans and low prevalence rate. Fully automated methods, with high accuracy, could therefore be useful. Various fully automated methods have been proposed, with the candidate detection step often using hand-crafted shape (Bian et al., 2013; Fazlollahi et al., 2014), intensity (Fazlollahi et al., 2015) and geometric features (Fazlollahi et al., 2014) within supervised classifier frameworks (Pan et al., 2008; Fazlollahi et al., 2014, 2015; Dou et al., 2015; Ghafaryasl et al., 2012). The FP reduction stage is typically based on supervised classifiers (Fazlollahi et al., 2015; Pan et al., 2008; Dou et al., 2015) using local intensity features and shape descriptors (e.g. Hessian-based shape descriptors (Fazlollahi et al., 2015)). Among the shape descriptors, the radial symmetry transform has been most commonly used (Bian et al., 2013; Liu et al., 2019b), exploiting the circular shape of CMBs. Hence, using structural (e.g. intensity and shape) and anatomical information in combination with the local characteristics (e.g. local contrast) could aid in the reduction of FPs and more accurate detection of CMBs (Dou et al., 2015).

Conventional machine learning (ML) methods require the extraction of meaningful features capable of distinguishing CMBs from the background and mimics. However, due to the small size and variation in shape and intensities of CMBs, designing robust, descriptive and cost effective features is highly challenging. The use of deep learning models, especially convolutional neural networks (CNNs) could overcome this challenge and provide more accurate CMB detection, since they efficiently extract both local and global contextual information. For instance, 3D CNN models have been used for feature extraction (Chen et al., 2015) and patch-level CMB detection (Dou et al., 2016). Dou et al. (2016) used a local region-based approach for segmentation of CMB candidates and discrimination of CMB and non-CMB patches. They initially trained a 3D CNN with true CMB samples and randomly selected background samples, applied the initial model on the training set and used the false positive patches for enlarging the training dataset in the discrimination step. Another region-based CNN method using You Only Look Once (YOLO) (Redmon and Farhadi, 2017) was proposed by Al-Masni et al. (2020) (using a 3D CNN was used for FP reduction). In addition to above methods, deep ResNets (He et al., 2016) were used for patch-level CMB classification (Chen et al., 2018; Liu et al., 2019b), along with a post-processing step using intensity morphological operations (Liu et al., 2019b). Given the size and sparsity of CMBs, class imbalance between CMBs and background is one of the major problems. Due to this, several methods used equal numbers of CMB patches and non-CMB patches, selected using manually annotated CMB voxels (and comparable number of non-CMB voxels) for training and evaluation purposes (Zhang et al., 2016, 2018; Wang et al., 2019; Hong et al., 2020; Lu et al., 2021b). Note that patches selected in these methods contained multiple CMBs, and therefore the patch-level performance might not correspond to the lesion-level performance. While the methods showed good performance at patch-level, the lesion-level performance is still more crucial to obtain clinically useful information (e.g. CMB location and count).

#### Knowledge distillation

Deep neural networks have been rapidly developing over recent years for accurate medical image segmentation tasks, including CMB segmentation, as mentioned above. However, the improved performance is achieved at the cost of long training times and using training resource-intensive complex models (Lan et al., 2018). Hence, training small networks, that are computationally efficient and generalisable across datasets, is highly desirable. With this aim of model compression (Bucilu et al., 2006), knowledge distillation (KD) (Hinton et al., 2015; Ba and Caruana, 2013) aims to train a smaller network (usually referred as *student network*) with the supervision (or distillation of knowledge) from a larger network (referred as *teacher network*). In KD, the student network is typically trained to match the prediction quality of the teacher network, and has been shown to reduce overfitting (Hinton et al., 2015; Lan et al., 2018). KD methods have been successfully used for various object detection tasks (Chen et al., 2017), including lesion segmentation on brain MR images (Lachinov et al., 2019; Vadacchino et al., 2021; Hu et al., 2020). The most commonly used distillation types include response-based (Hinton et al., 2015; Kim and Kim, 2017; Müller et al., 2019; Ding et al., 2019) and feature-based distillation (Romero et al., 2014; Jin et al., 2019; Zhou et al., 2018). In response-based distillation, the output logits from the softmax layer are softened (also known as *soft labels*) using a *temperature* parameter that acts as a regularisation factor (Hinton et al., 2015). In the feature-based distillation, outputs of intermediate layers of the teacher model are used to train the student model (e.g. *hint learning* using outputs of hidden layers (Jin et al., 2019; Romero et al., 2014) and parameter sharing of intermediate layers (Zhou et al., 2018)).

Based on the training methods, offline distillation (using a pretrained teacher model to train the student model) (Hinton et al., 2015; Romero et al., 2014), online distillation (training teacher and student models together) (Guo et al., 2020; Zhou et al., 2018) and self-distillation (where the student model from prior epochs becomes the teacher for the subsequent epochs) (Zhang et al., 2019; Yang et al., 2019) are most commonly used. Various techniques have also been proposed to improve the generalisability and the performance of the student models including using noisy data (Li et al., 2017; Sarfraz et al., 2019), adaptive regularisation of distillation parameters (Ding et al., 2019) and adversarial perturbation of data for training (Xie et al., 2020). Multi-task learning methods have also been shown to provide a good regularisation, reducing the risk of over-fitting (Liu et al., 2019a; Ye et al., 2019). The auxiliary task could be a related task (e.g. auxiliary classification network in lesion segmentation (Yang et al., 2017)) or an adversarial task (e.g. adversarial training of domain predictor in domain adaptation networks (Ganin et al., 2016)).

So far, KD has never been used for CMB detection to the best of our knowledge. However, a complex teacher network that captures the subtle differences in the structural pattern of CMBs would be highly beneficial to efficiently enhance the ability of a smaller student model in differentiating CMBs not only from the healthy tissue, but also from mimics. In this work we use a knowledge distillation framework, for the first time, for accurate and fully automated detection of CMBs in a computationally light manner. We propose a 2-step CMB detection - CMB candidate detection and discrimination of CMB candidates into CMBs or non-CMBs - using 3D CNN models in both steps. We tested our approach in the presence of mimics, across different datasets with different modalities and pathological conditions. Our main contributions are as follows:

- In the initial CMB candidate detection step (section 2.2), we utilise the radial symmetry property of CMBs for more efficient and accurate candidate detection.
- In the candidate discrimination step (section 2.3), we use a knowledge distillation framework to create a light-weight student model from a multi-tasking teacher model, which overcomes the class imbalance between CMBs and the background, leading to effective removal of false positives.
- We evaluated our method on 4 different datasets. The dataset details are provided in section 3. Through the experiments described in section 4, we studied the contribution of the individual steps on the CMB detection performance, and also the effect of various modalities and different pathological conditions on the detection results.

We also performed an indirect comparison of our results with existing methods at various stages of detection.

## 2. CMB detection method

### 2.1. Data preprocessing

We reoriented the T2*-GRE, SWI and QSM images to match the orientation of the standard MNI template, and skull stripped the images using FSL BET (Smith, 2002). For T2*-GRE and SWI, we performed bias field correction using FSL FAST (Zhang et al., 2001). We also inverted the intensity values of the input volume by subtracting the intensity-normalised image (obtained by dividing intensity values by the maximum intensity) from 1, so that CMBs have higher intensities (to facilitate our choice of CNN layers). For QSM images, we only normalised the intensity values without inverting their intensity values since CMBs already appear hyperintense with respect to the background. We cropped the skull stripped images closer to the brain edges to make the FOV tighter.

#### 2.1.1. Removal of blood vessels and sulci

In the first step, we aim to remove the blood vessels, sulci and other elongated structures in the input image to reduce the appearance of CMB mimics. For the removal of blood vessels and sulci, we used the method described in Sundaresan et al. (2021). Briefly, the method involves the extraction of edge and orientation-based features, using Frangi filters (Frangi et al., 1998) and eigenvalues of the structure tensor (Förstner, 1994), followed by K-means clustering to obtain the vessel mask. The masked regions were then inpainted using the mean of intensity values from the immediate non-masked neighbouring voxels (within a 26-connected neighbourhood). Figure 1 shows a few sample images (from various modalities) after removal of vessels and sulci.

**Figure 1:**
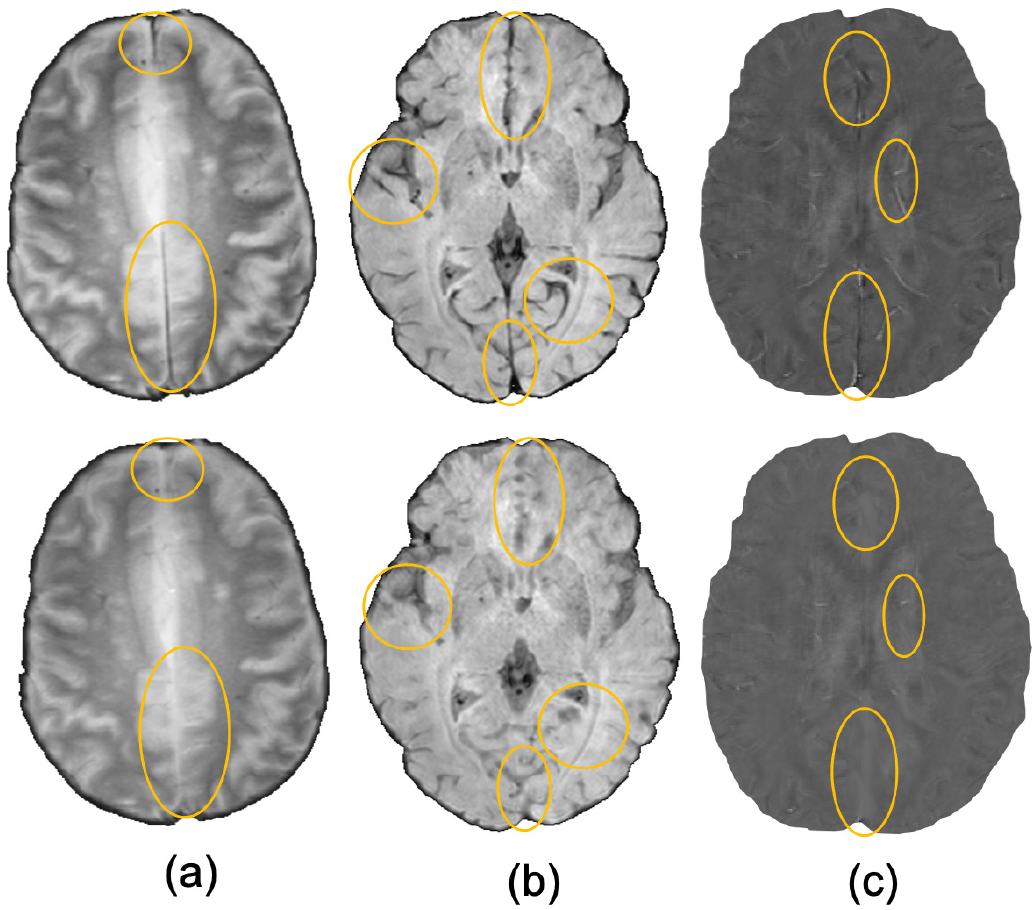
Sample images after removal of blood vessels and sulci (regions with major changes indicated by circles in the bottom row) shown for (a) T2*-GRE, (b) SWI and (c) QSM modalities.

### 2.2. 3D CMB initial candidate detection

For the initial CMB candidate detection, in addition to the intensity characteristics, we also use the radial symmetry property of CMBs. Hence, we performed a fast radial symmetry transform (FRST) (Loy and Zelinsky, 2002) with four radii of 2, 3, 4 and 6 voxels. We computed the mean of the outputs at the above radii to obtain the final FRST output (shown for different modalities in figure 2). For both the input modality and the FRST output, we split the 3D volumes into patches of size 48 × 48 × 48 voxels and provided them as 2 input channels to the 3D patch-based encoder-decoder model for initial candidate detection. We selected the patch-size of 48 voxels empirically - at this scale, the patches were large enough to overcome the effect of local noise and assign higher probabilities to CMB-like regions on experimented datasets described in section 3.

**Figure 2:**
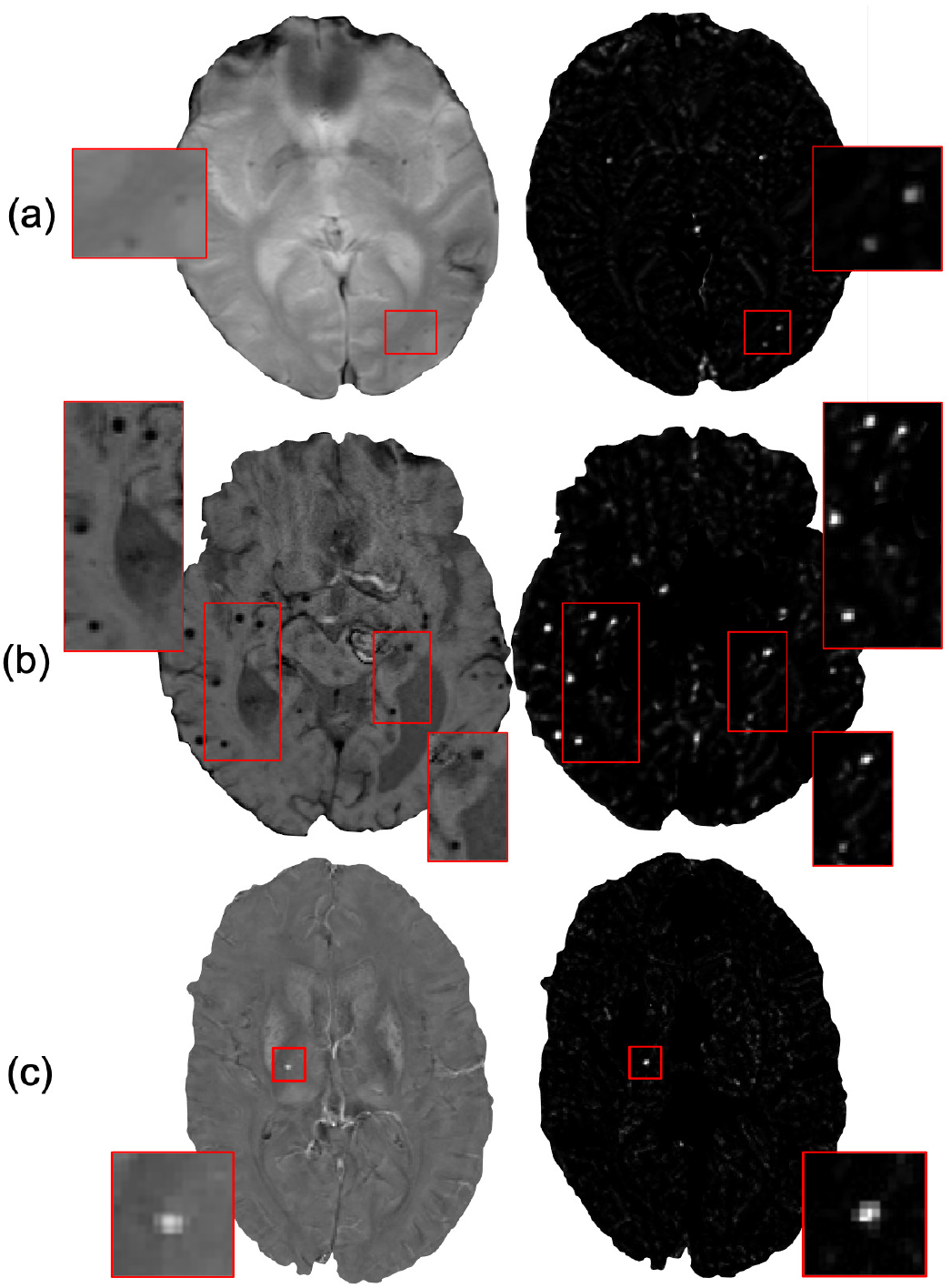
Examples of the final fast radial symmetry transform (FRST) outputs for various image modalities. The FRST outputs shown (in the right panel) for (a) T2*-GRE, (b) SWI and (c) QSM images. Inset figures show the magnified versions of the regions indicated in the boxes.

Figure 3 shows the block diagram of this step and the architecture of the 3D encoder-decoder model used in the initial candidate detection. The architecture of the 3D encoder-decoder network at a scale *N* is based on a shallow U-Net. We trimmed the U-Net to a shallow architecture with 2 pooling layers, since CMBs are small and depend more on local neighbourhood, rather than the global context. The input channels are converted into 3 channels by the initial 1 × 1 × 1 projection layer, followed by 3 × 3 × 3 convolution to get the initial filter channel depth of 64. The architecture consists of two consecutive 3 × 3 × 3 convolutional layers followed by the 2 × 2 × 2 max-pooling layer (in the case of encoder) or 2 × 2 × 2 upsampling layer (in the case of decoder). We added a 1 × 1 × 1 convolutional layer before the final softmax layer for predicting the probability maps *P*_*Cdet*_.

**Figure 3:**
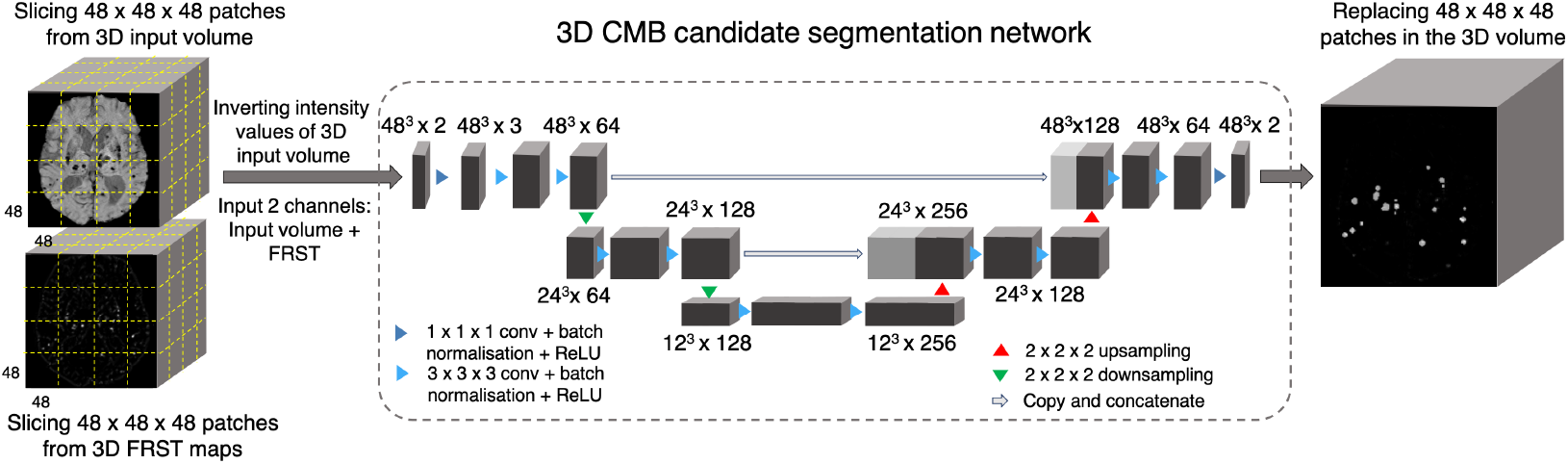
CMB initial candidate detection step. Block diagram of the candidate detection step with the architecture of the 3D encoder-decoder network based on shallow U-Net.

#### 2.2.1. Loss functions

We used a combination of cross-entropy (CE) and Dice loss functions as the total loss function. In the CE loss function, we upweight the CMB voxels 10 times compared to the non-CMB voxels during training to compensate for the imbalance in the classes. Dice loss is based on the voxel-wise Dice similarity measure and aids in the accurate detection of edges and small CMBs in the patches.

### 2.3. CMB candidate discrimination

The candidate discrimination step is more challenging than the initial candidate detection step, since the discrimination step needs to learn the subtle features to detect CMBs and discriminate them from other CMB mimics. To illustrate the complexity of the problem, figure 4 shows instances of CMB and non CMB patches that were all identified as CMB candidates in the initial detection step. In this step, we use a student-teacher framework for classifying true CMB candidates from FPs. We use two networks: (1) a teacher network that has a more complex architecture (and hence is computationally expensive) and learns the task-based characteristics (in our case, CMB-related features) from a larger dataset; (2) a student model that has comparatively simpler architecture, that enables faster training on different datasets, and is trained directly on the patches centred at candidates detected from the initial candidate detection step (section 2.2). We aim to improve the classification accuracy of the student model, by guiding its training using the teacher model with response-based knowledge distillation. Figure 5 shows the proposed overall architecture and details of the student-teacher architecture and training are provided in the sections below.

**Figure 4:**
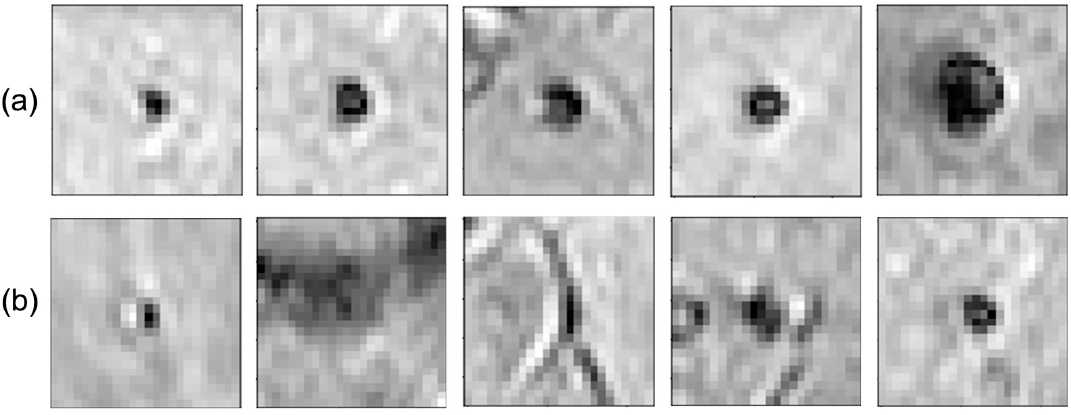
Examples of initial candidates detected in the first step. (a) CMB and (b) non-CMB patches are shown separately. Note that in most of the cases, non-CMB instances are quite similar to CMBs.

**Figure 5:**
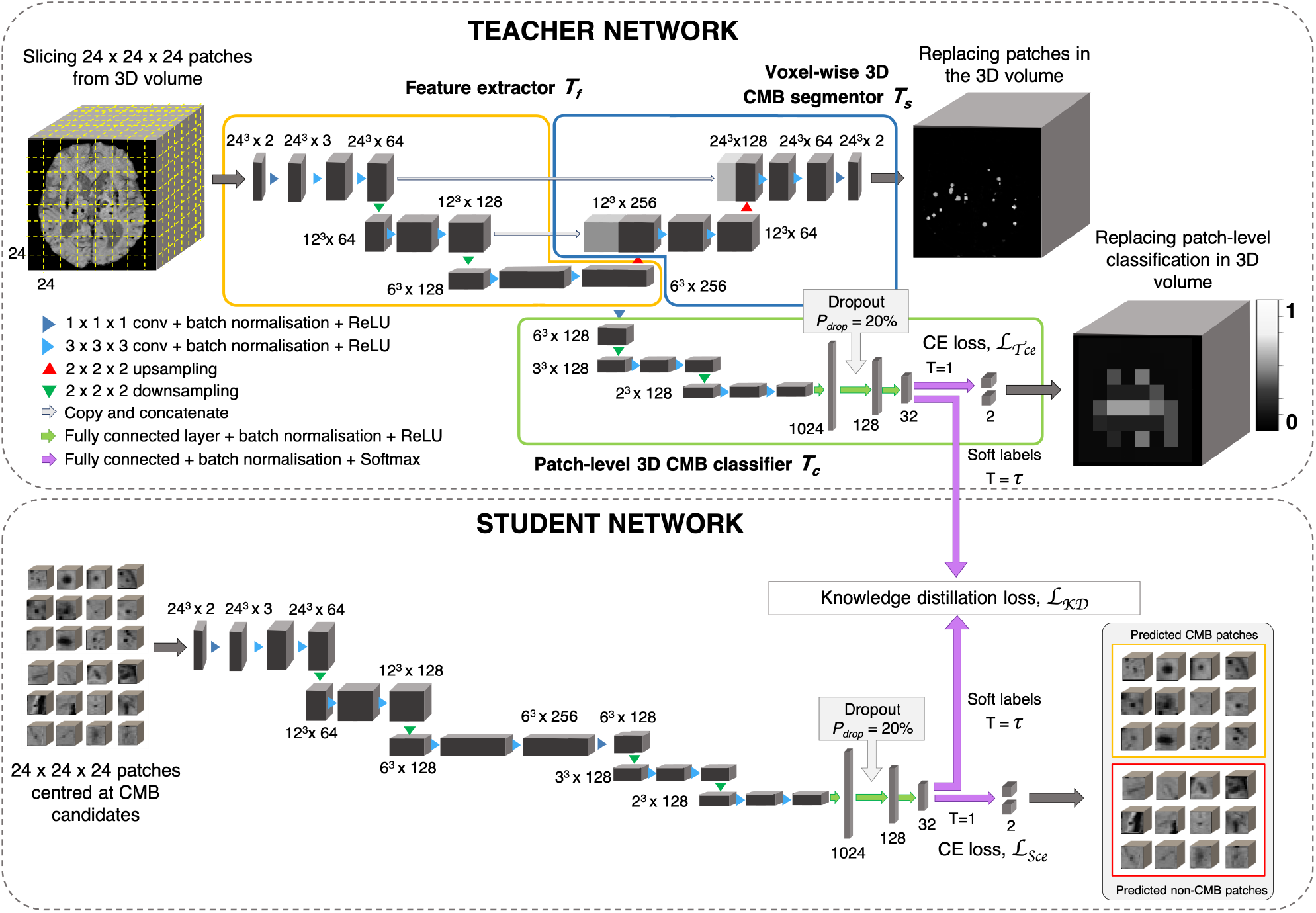
CMB candidate discrimination using knowledge distillation. Top panel shows the multi-tasking teacher model consisting of feature extractor *T*_*f*_ (orange), voxel-wise 3D CMB segmentor *T*_*s*_ (blue) and patch-level 3D CMB classifier *T*_*c*_ (green). The bottom panel shows the student model *S* for classification of CMB and non-CMB patches using the distillation of knowledge from the teacher model using the distillation loss *L*_*KD*_(*T*_*c*_, *S*).

#### 2.3.1. Teacher network with multi-task training

The teacher model uses a multi-tasking architecture consisting of three parts (1) feature extractor (*T*_*f*_), (2) voxel-wise CMB segmentor (*T*_*s*_) and (3) patch-level CMB classifier (*T*_*c*_). We took the pretrained network used for the initial CMB candidate detection and added a patch-level CMB classifier arm to the model. While the architecture of feature extractor + segmentor is the same as that of the model used in the initial candidate detection stage, the classifier arm consists of a projection layer with 1 × 1 × 1 kernel, followed by two consecutive 3 × 3 × 3 convolutional layers followed by a pooling layer in each level of abstraction. The output of the third layer of the encoder is fed into dense fully connected layers (FC). Three fully connected layers (FC-1024, FC-128 and FC-32) with 1024, 128 and 32 nodes are then followed by a softmax layer. We added a dropout layer with *drop probabilities* of 20% before the FC-128 layer. We extracted 24 × 24 × 24 adjacent patches from the input modality and FRST images, and provide them as 2-channeled input for training this model. While we used a patch-size of 48 for the detection stage, we used a smaller patch-size of 24 for this stage. This is because our main aim was to fine-tune the pretrained segmentor model to determine the lesion-level characteristics of CMBs from the local neighbourhood given the initial candidates, and use this contextual information to train the patch-level CMB classifier arm. The *T*_*f*_, made of a series of convolutional layers, extracts features that are helpful for both *T*_*s*_ and *T*_*c*_. Therefore, both *T*_*s*_ and *T*_*c*_ learn to improve the CMB segmentation and classification in a progressive manner since both are trained simultaneously with shared weights in *T*_*f*_. This means that *T*_*s*_ assigns high probability values to the CMB voxels in the CMB patches, while reducing the probability values of CMB-like mimics on the non-CMB patches. At the same time, *T*_*c*_ detects the patches with more CMB-like features (regions that are assigned higher probabilities by *T*_*s*_) as CMB patches with higher confidence and vice versa. In addition to the loss function to train *T*_*s*_ (specified in section 2.2.1), we used a binary cross-entropy loss function for *T*_*c*_.

#### 2.3.2. Knowledge distillation using student network

The student model consists of the feature extractor and patch-level classifier parts (*T*_*f*_ + *T*_*c*_) of the teacher model. We trained the student model in an offline manner using response-based knowledge distillation (KD). While we provided adjacent 24 × 24 × 24 patches for the teacher model, we extracted more meaningful input patches for the student model, centred at the detected initial CMB candidates for quicker learning. For determining the centroids, we thresholded *P*_*Cdet*_ from the first step at a specific threshold *Th*_*Cdet*_ based on the performance values (for more details refer to section 5.2). During testing, patches centred at candidates detected from the initial candidate detection step are classified as CMB or non-CMB by the student model. Let the student model and teacher model classifier be *S* and *T*_*c*_ respectively. For the distillation of knowledge from the teacher model for training the student model, the loss function is given by

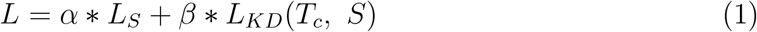

where *L*_*s*_ is the student loss function, *L*_*KD*_(*T*_*c*_, *S*) is the KD loss and *α, β* are weighing parameters. We used the cross-entropy loss function as the student loss. For determining the KD loss, the targets are the class outputs predicted by the classifier of the teacher model (in the inference mode) on the same input as that of the student model. A temperature (*τ*) parameter is used in the softmax function to soften the target distribution. While *τ* = 1 provides the usual softmax outputs, higher values of *τ* softens the softmax outputs (as shown in eqn. 2). The softmax function with *τ* is given by,

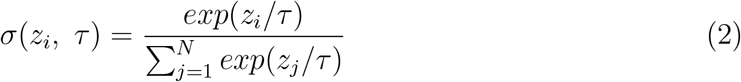

where N is the number of classes. Compared to hard target distributions (closer to 0 or 1 for individual classes), softer target distributions (between 0 and 1) have been shown to aid in training generalisable student model (Hinton et al., 2015), however having very high *τ* might also be counter-productive in some cases. The optimal value of *τ* and the level of softness in the target distribution depends on specific applications, student/teacher network architectures and dataset characteristics. Temperature *τ* values between 2.5 and 4 have been shown to provide better results, while models with more units in the hidden layers may require higher *τ* values (Hinton et al., 2015). Using the temperature *τ* parameter, the KD loss is given by,

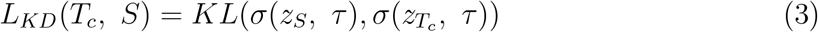

where *KL* is the KL-divergence (distance between the class probability distributions of student and teacher classifier models). From eqn. 1, the loss function is,

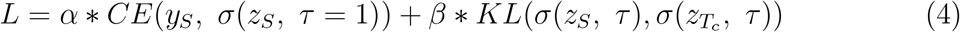

where *y*_*S*_ are the target labels of the student model and *z*_*S*_ and 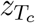 are the logits (inputs to the softmax layer) of student and teacher classifier model respectively.

#### 2.3.3. Post-processing

We applied a threshold *Th*_*Cdisc*_ on the patch-wise probabilities to discriminate CMB and non-CMB candidates. We set *Th*_*Cdisc*_ values empirically based on the performance metric values (refer to section 5.2). Additionally, we removed the noisy stray voxels by filtering out the candidates with volume *<* 5 voxels, removed the tubular structures (e.g. fragments of sulci near the skull) by filtering out candidates having higher ellipticity (*>* 0.2) and removed the CMB candidates that are closer to skull (*<* 5 voxels from the brain mask boundary) to reject the FPs due to the sulci in the brain. The above values were determined empirically based on the results on an independent dataset, as specified in section 2.5.

### 2.4. Data augmentation

Due to the small size of CMBs, transformations such as rotation and down-scaling could result in the loss of CMBs in the augmented data patches. Hence, we chose our data augmentation carefully, to inject variations in the data with minimal interpolation of intensity values. For the initial candidate detection step, we performed augmentation on the patches, increasing the dataset size by a factor of 10, using random combinations of the following transformations: translation, random noise injection and Gaussian filtering (with a small *σ* value). The parameters for the above transformations were chosen randomly from the ranges reported in table 1. We used similar augmentation for the discrimination step, increasing training data by a factor of 5.

**Table 1:** Transformations used for data augmentation and their parameter ranges.

| Transformations |  | Parameter ranges |
| --- | --- | --- |
| Translation | | x-offset: $[-15, 15]$, y-offset: $[-15, 15]$ voxels |
| Random noise injection | | Distribution: Gaussian, $\mu = 0$, $\sigma^2 = [0.01, 0.04]$ |
| Gaussian filtering | | $\sigma = [0.1, 0.2]$ voxels |

### 2.5. Implementation details

For both CMB candidate detection and discrimination steps, we trained the networks using the Adam Optimiser (Kingma and Ba, 2014) with epsilon (*c*) value of 1 × 10^−4^. We used a batch size of 8, with an initial learning rate of 1 × 10^−3^ and reducing it by a factor 1 × 10^−1^ every 2 epochs, until it reaches 1 × 10^−6^, after which we maintain the fixed learning rate value. For both candidate detection and candidate discrimination training, we also empirically set the total number of epochs to 100 and used a criterion based on a patience value (number of epochs to wait for progress on validation set) of 20 epochs to determine model convergence for early stopping. We used the truncated normal variable (with *σ* = 0.05) for weights initialisation and the biases were initialised as constants with a value of 0.1. For the student model in the discrimination step, we used a temperature *τ* = 4, *α* = 0.4 and *β* = 0.6. We used a subset of a publicly available dataset (https://valdo.grand-challenge.org/Data/) consisting of a random sample of 20 subjects for hyper-parameter tuning and the empirical determination of parameter values in the post-processing step and loss functions. Our experiments were completed on NVIDIA Tesla V100 GPU and were implemented in Python 3.6 using Pytorch 1.2.0.

## 3. Datasets used

We used following datasets for the evaluation of our proposed method. The datasets consist of images from different modalities, that were acquired using different scanners with variations in acquisition protocols and from subjects with different pathological conditions and demographic characteristics.

### 3.1. The UK Biobank (UKBB) dataset

From 14,521 subjects (out of *≈* 20,000 subjects from the January 2018 release of UKBB), we preselected 78 CMB candidate subjects using the method proposed in Sundaresan et al. (2021). Manual segmentations in the form of coordinates were annotated on SWI images for these 78 subjects by a trained radiologist (A.G.M). From those coordinates, the ground truth segmentation for each CMB was obtained by a region-growing-based method that in addition to a voxel’s intensity also takes into account its distance from the seed voxel, and is constrained by a maximum radius of 5 voxels in-plane and 3 voxels through-plane. The age range of subjects is 50.8 - 74.8 years, mean age 59.9 *±* 7.2 years, median age 57.8 years, female to male ratio F:M = 37:41. For SWI, 3D multi-echo GRE images were acquired using 3T Siemens Magnetom Skyra scanner with TR/TE = 27/9.4/20 ms, flip angle 15^*o*^, voxel resolution of 0.8 × 0.8 × 3 mm, with image dimension of 256 × 288 × 48 voxels. The QSM images were generated using a multi-step post-processing of phase data as described in Wang et al. (2021). Briefly, the method involved combination of phase data of individual channels, phase unwrapping, background field removal, followed by dipole inversion. Total number of CMBs in this dataset: 186, mean: 2.4 *±* 7.0 CMBs/subject, median: 1 CMB/subject.

### 3.2. The Oxford Vascular Study (OXVASC) dataset

The dataset consists of T2*-GRE images from 74 participants from the OXVASC study (Rothwell et al., 2004), who had recently experienced a minor non-disabling stroke or transient ischemic attack. The 2D single-echo T2*-GRE images were acquired using 3T Siemens Verio scanner with GRAPPA factor = 2, TR/TE = 504/15 ms, flip angle 20^*o*^, voxel resolution of 0.9 × 0.8 × 5 mm, with image dimension of 640 × 640 × 25 voxels. Age range 39.6 - 91.2 years, mean age 69.8 *±* 14.6 years, median age 67.3 years, female to male ratio F:M = 36:38. Out of 74 subjects, 36 subjects had CMBs, and manual segmentations, labelled using T2*-GRE images, were available for all 36 subjects. Total number of CMBs: 366, mean: 10.2 *±* 33.3 CMBs/subject, median: 3 CMBs/subject.

### 3.3. The Tranexamic acid for IntraCerebral Haemorrhage 2 (TICH2) trial MRI sub-study dataset

The dataset consists of a subset of the MRI data used in Dineen et al. (2018) obtained as part of the TICH2 trial (Sprigg et al., 2018). The dataset consists of images with variations in image dimension, spatial resolution and MR acquisition parameters (details in Dineen et al. (2018)). The dataset used in this work consists of 115 SWI from the subjects with spontaneous intracerebral haemorrhage (ICH). Age range 29 - 88 years, mean age 64.76 *±* 15.5 years, median age 66.5 years, female to male ratio F:M = 24:26. Out of 115 subjects, 71 subjects had CMBs and manual segmentations for CMBs were available for all 71 subjects. Additionally, microbleed anatomical rating scale (MARS, Gregoire et al. (2009)) values were provided for the CMB subjects. For evaluation purposes we included in the manual segmentation maps used in all our experiments all CMBs that were labelled as either ‘*definite*’ or ‘*possible*’. Total number of CMBs: 849, mean: 11.9 *±* 22.0 CMBs/subject, median: 3 CMBs/subject.

### 3.4. The stroke dataset from Hong Kong (SHK)

Originally, the dataset used in Dou et al. (2016) consisted of 320 SWI images in total, out of which 126 are subjects with stroke (mean age: 67.4 *±* 11.3) and 194 are from normal ageing subjects (mean age: 71.2 *±* 5.0). In this work, we used a subset of 20 subjects that were publicly available from this dataset. Manual annotations in the form of CMB coordinates were available along with the dataset. From coordinates, ground truth segmentations were obtained with the method used for the UKBB data (refer section 3.1). Another rater independently provided the manual segmentations on SWI images on the dataset, and we considered the union of both manual masks as our final ground truth. Total number of CMBs: 126, mean: 6.3 *±* 8.8 CMBs/subject, median: 3 CMBs/subject.

## 4. Experiments

### 4.1. Performance evaluation metrics

We evaluated the CMB detection results using the following metrics for total number of CMBs over the individual datasets, as done in the existing literature:

- **Cluster-level TPR:** the number of true positive clusters (i.e. CMBs) divided by the total number of true clusters as given by,

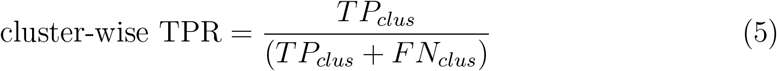

where *TP*_*clus*_ and *FN*_*clus*_ are true positive (overlaps with a ground truth cluster by at least one voxel) and false negative clusters respectively.
- **Average number of FPs per subject (FPavg):** for a given dataset *D*, FPavg is defined as the ratio of the total number of detected FP clusters (*FP*_*clus*_, has no overlap with a ground truth cluster) to the number of subjects (or images) in the dataset, as given by,

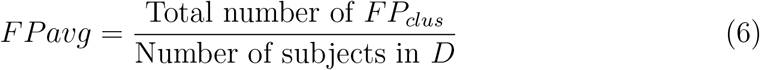
- **Cluster-wise precision:** the number of true positive clusters divided by the total number of detected clusters as given by,

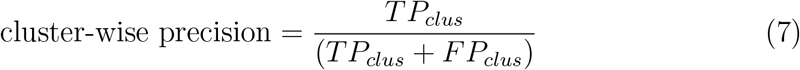

We used 26-connectivity to form the clusters. We used TPR and FPavg values for plotting a free-response receiver operating characteristics (FROC) curve, which is a plot of cluster-wise TPR versus the average number of false positives per image/subject.

### 4.2. Ablation study: effect of knowledge distillation on CMB detection

In this study, we evaluate the effect of individual steps, including the teacher-student distillation framework and the post-processing step on the CMB detection performance (using metrics specified in section 4.1) on the UKBB dataset (using a training-validation-test split of 44-10-24 subjects, with 40 CMBs in the test data). To this aim, we obtain the above performance evaluation metrics at the following stages: (i) after the initial CMB candidate detection, (ii) after candidate discrimination using a classification network trained without the teacher model (trained independently using only CE loss function *L*_*S*_), (iii) after candidate discrimination with knowledge distillation using student-teacher training and (iv) after final post-processing.

### 4.3. Cross-validation of CMB detection on T2*-GRE and SWI images

We performed 5-fold cross-validation separately on T2*-GRE images from the OXVASC dataset and SWI images from the UKBB dataset, and evaluated the cluster-wise performance using the metrics specified in section 4.1. Note that for this cross-validation, we used the hyper-parameters that were determined separately using an independent dataset specified in section 2.5.

### 4.4. Evaluation of the generalisability of the proposed method

We trained the proposed method on SWI images from 78 subjects from the UKBB dataset for this experiment (using the hyperparameters mentioned in section 2.5) and evaluated the method on data from different domains (e.g. variations in intensity profiles, scanners and acquisition protocols and demographics), using performance metrics specified in section 4.1, under the following three scenarios:

#### Evaluation on the same dataset with different modality

We evaluated the effect of change in the modality only on CMB detection by applying the method, that was trained on SWI images from the UKBB dataset, to the QSM images from the same subjects from the UKBB dataset.

#### Evaluation on different datasets with same modality

We evaluated our method on different test datasets to observe the effect of scanner-related and population-level pathological variations on the CMB detection. We applied our method trained on SWI images from the UKBB dataset to SWI images from 115 subjects with intra-cerebral haemorrhages from the TICH2 dataset and SWI images from 10 healthy controls and 10 subjects with stroke from the SHK dataset.

#### Evaluation on different datasets with different modality

We evaluated our method trained on SWI images from the UKBB dataset on the T2*-GRE images from 74 subjects from the OXVASC dataset. The OXVASC data is quite different from the UKBB data not only in terms of modality, but also in terms of resolution, scanner and demographic/pathological factors. Hence, this scenario would provide a better indication of the method’s generalisability in real world clinical applications.

For the above experiments, for the CMB candidate detection and discrimination steps, we used the threshold values (*Th*_*Cdet*_ and *Th*_*Cdisc*_) determined during 5-fold cross-validation on the UKBB dataset.

### 4.5. Comparison of our results with the existing literature

Finally, we performed an indirect comparison of our results from the UKBB and OXVASC datasets with those of existing CMB detection methods in the literature.

## 5. Results

### 5.1. Ablation study: effect of knowledge distillation on CMB detection

Figure 6 shows the FROC curves for the initial CMB candidate detection and candidate discrimination steps of our method on the UKBB dataset. Table 2 reports the best performance points at the ‘knee-point’ on the FROC curves for the 2 steps, along with the performance metrics after post-processing. In the candidate detection step, the aim was to achieve higher cluster-wise TPR, to detect as many true CMBs as possible. Hence, the number of FPs were higher at this step (with the highest cluster-wise TPR of 0.975 at the *Th*_*Cdet*_ = 0.5), when compared to the subsequent steps. For the candidate discrimination step (figure 6b), the FROC curves are shown for the comparison of the student network trained with KD framework from the teacher model and classification network (with same architecture as that of the student network) trained independently without KD. The performance at the candidate discrimination step is better with KD (cluster-wise TPR of 0.9 at *Th*_*Cdisc*_ = 0.3) than the model trained without KD (cluster-wise TPR of 0.75 at *Th*_*Cdisc*_ = 0.4), with the former showing an improvement of 0.02 in the cluster-wise precision (see table 2).

**Table 2:**
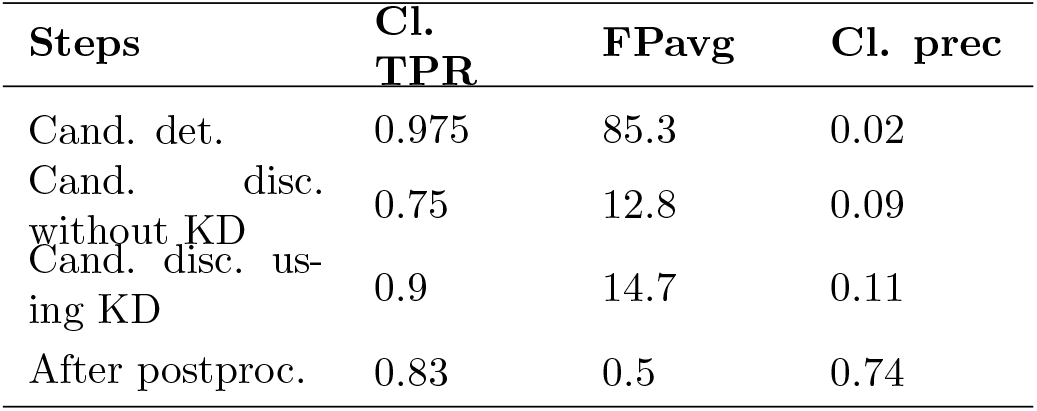
Ablation study: performance metrics after candidate detection, discrimination and postprocessing steps. Cl. TPR and Cl. prec indicate cluster-wise TPR and cluster-wise precision respectively.

**Figure 6:**
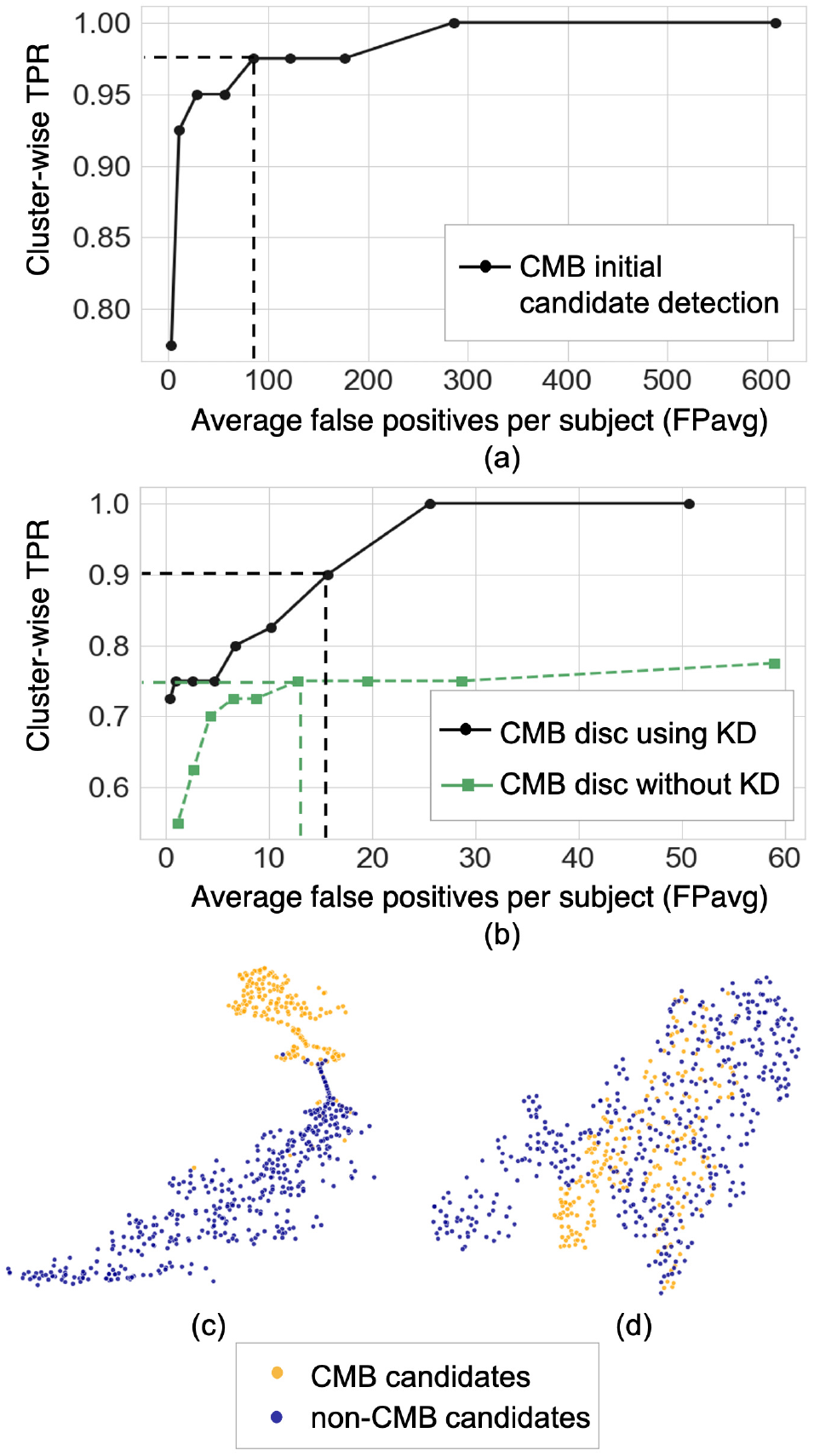
Results of the ablation study. (a) FROC curves at the CMB initial candidate detection stage, (b) FROC curves comparing the classification performance of the student model trained using KD from a teacher model (black solid •) and the same model trained independently without KD (green dashed ▪). The horizontal and vertical dashed lines on the FROC curves indicate the best performance points (reported in table 2) at *Th*_*Cdet*_ = 0.5 at the candidate detection step, *Th*_*Cdisc*_ = 0.3 and 0.4 for the models with and without KD respectively in the candidate discrimination step. T-SNE plots showing feature embeddings at the FC-32 layer for CMB (orange) and non-CMB (dark blue) cases for (c) the student model trained using KD and (d) the model without using KD.

The t-stochastic neighbor embedding (t-SNE) plots in figure 6c and 6d show the feature embeddings of the last fully connected layer (FC-32) in the classification network (in the CMB discrimination step) for the CMB and non-CMB cases, trained with and without KD. The feature embeddings for the student model using KD were quite separable between the CMB and non-CMB cases, indicating the capability of the student model to learn the subtle differences in the features between CMB and non-CMB classes. The classification model without KD, on the other hand, showed substantially more overlap between the feature embeddings. The postprocessing step improves the cluster-wise precision. Upon visual inspection, the main reductions in FPs were near the skull (e.g. sulci), penetrating blood vessels and stray noisy voxels. Regarding the contribution of individual attributes (e.g. shape, area and proximity to the skull) in FP reduction, we observed that around 65%, 25% and 15% of FPs after applying thresholds on distance of candidates from the skull, area and shape of candidates successively. Since the individual thresholds were determined as a part of hyper-parameter tuning on an independent dataset (section 2.3.3) and the interaction between the three attributes’ thresholds on FPavg is difficult to visualise, a separate FROC curve for the post-processing step is not shown. The majority of FPs rejected at this stage consists of candidates closer to the skull - these candidates passed the discrimination step since most of the CMBs in the training data (for the student model) were lobar CMBs and were closer to skull. Hence the discrimination step (despite removing a large amount of FPs near sulci) allows false predictions in this region. Having said that, it is worth noting that, in the post-processing step, a few true CMBs closer to the skull were also rejected as FPs, hence leading to a slight decrease in the cluster-wise TPR values.

### 5.2. Cross-validation of CMB detection on T2*-GRE and SWI images

Figure 7 shows the FROC curves for CMB candidate detection and candidate discrimination steps of 5-fold cross-validation on the UKBB and OXVASC datasets. Table 3 reports the best performance metrics at different steps of the cross-validation on the UKBB and OXVASC datasets. The proposed method achieved cluster-wise TPR values of 0.93 and 0.90 with FPavg of 1.5 and 0.9 at *Th*_*Cdet*_, *Th*_*Cdisc*_ = 0.3 and 0.2 on the UKBB and OXVASC datasets respectively. The method provides higher cluster-wise TPR and FPavg values on SWI images (from the UKBB dataset) when compared to the T2*-GRE images from the OXVASC dataset. Even though the FPavg values were comparable at the candidate detection step for both datasets, the student model at candidate discrimination step provided much lower FPavg on T2*-GRE images from the OXVASC dataset, thus providing higher cluster-wise precision value. The FPavg values reduced substantially after the post-processing step with only slight reduction in the cluster-wise TPR values.

**Table 3:**
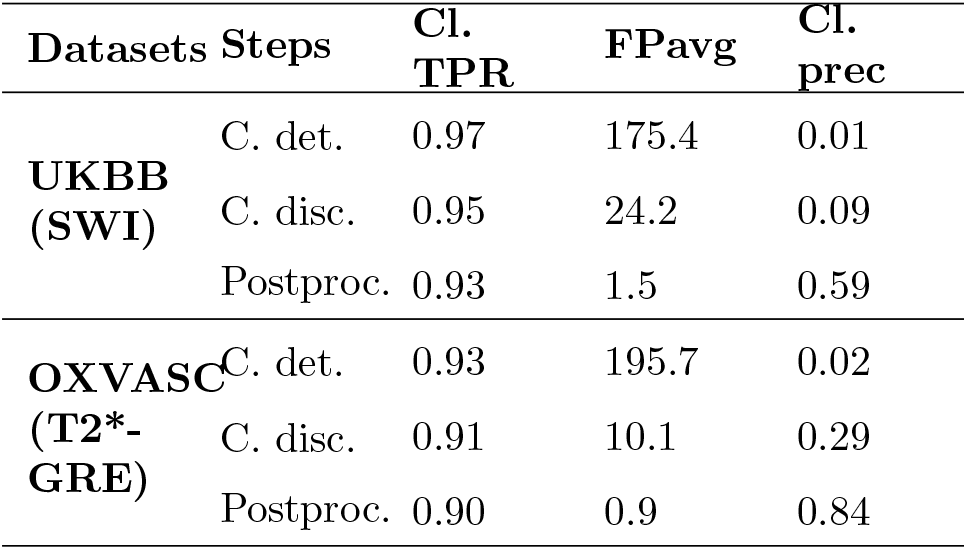
Cross-validation on the UKBB and OXVASC datasets: performance metrics at candidate detection, discrimination and post-processing steps. Cl. TPR and Cl. prec indicate cluster-wise TPR and cluster-wise precision respectively. C. det - candidate detection, C. disc - candidate discrimination.

**Figure 7:**
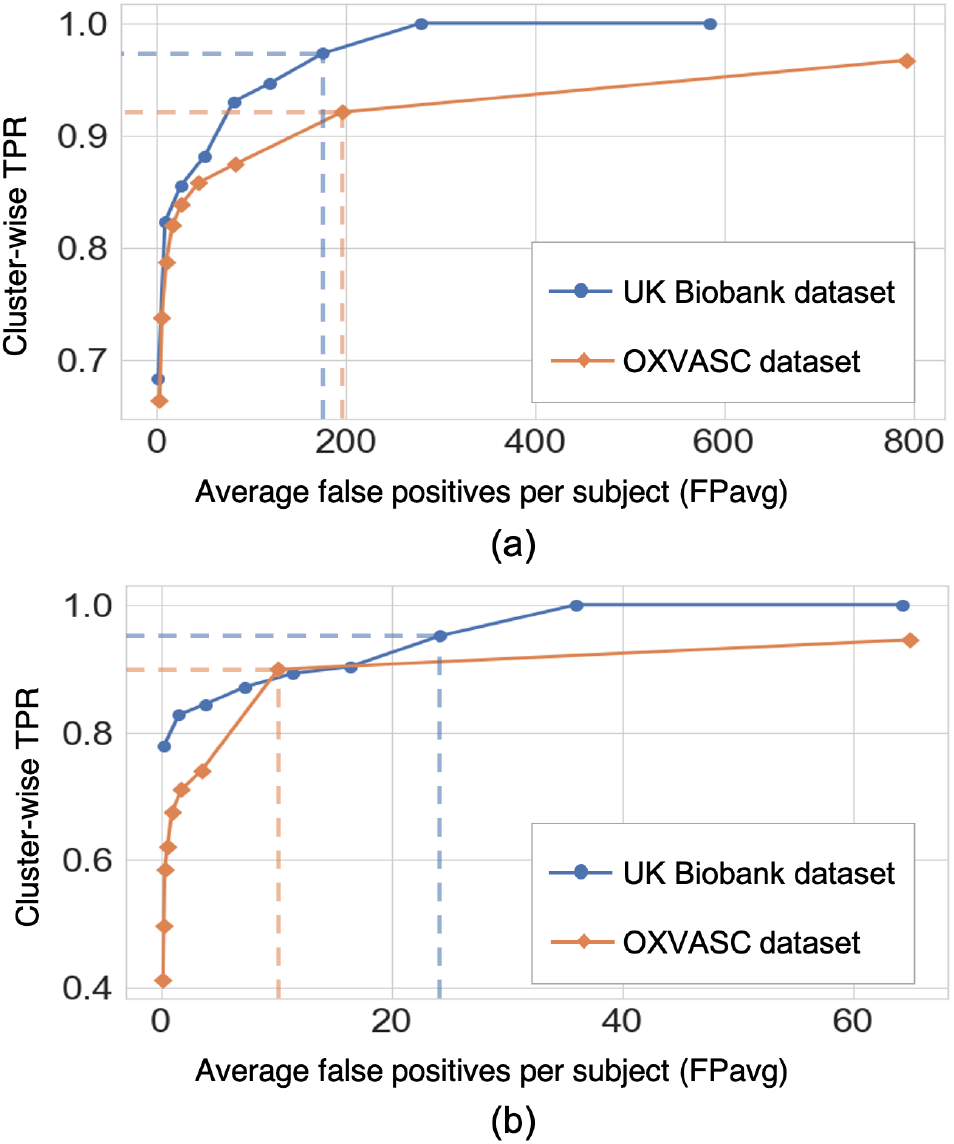
Results of the 5-fold cross-validation. FROC curves at (a) the CMB initial candidate detection stage and (b) the candidate discrimination stage on the UKBB (blue) and OXVASC (orange) datasets. The dashed lines on the FROC curves indicate the best performance points (reported in table 3) at specific threshold points (threshold = 0.3 and 0.2 for the UKBB and OXVASC datasets respectively for both candidate detection and candidate discrimination steps.)

Figure 8 shows sample results of the cross-validation at various steps of CMB detection on the UKBB and the OXVASC datasets. In both UKBB and OXVASC datasets, the main sources of FPs in the initial candidate detection step are sulci, minor intensity/contrast variations in the brain tissue and small vessel fragments. While most of the penetrating blood vessels are segmented correctly as part of the background even at the candidate detection step (due to the vessel removal step, especially in the OXVASC dataset), the remaining FPs on/near the vessels are removed at the discrimination step. The post-processing step further reduced the stray noisy voxels (with volume *<* 5 voxels) and sulci regions closer to the skull, resulting in very few FPs on both datasets.

**Figure 8:**
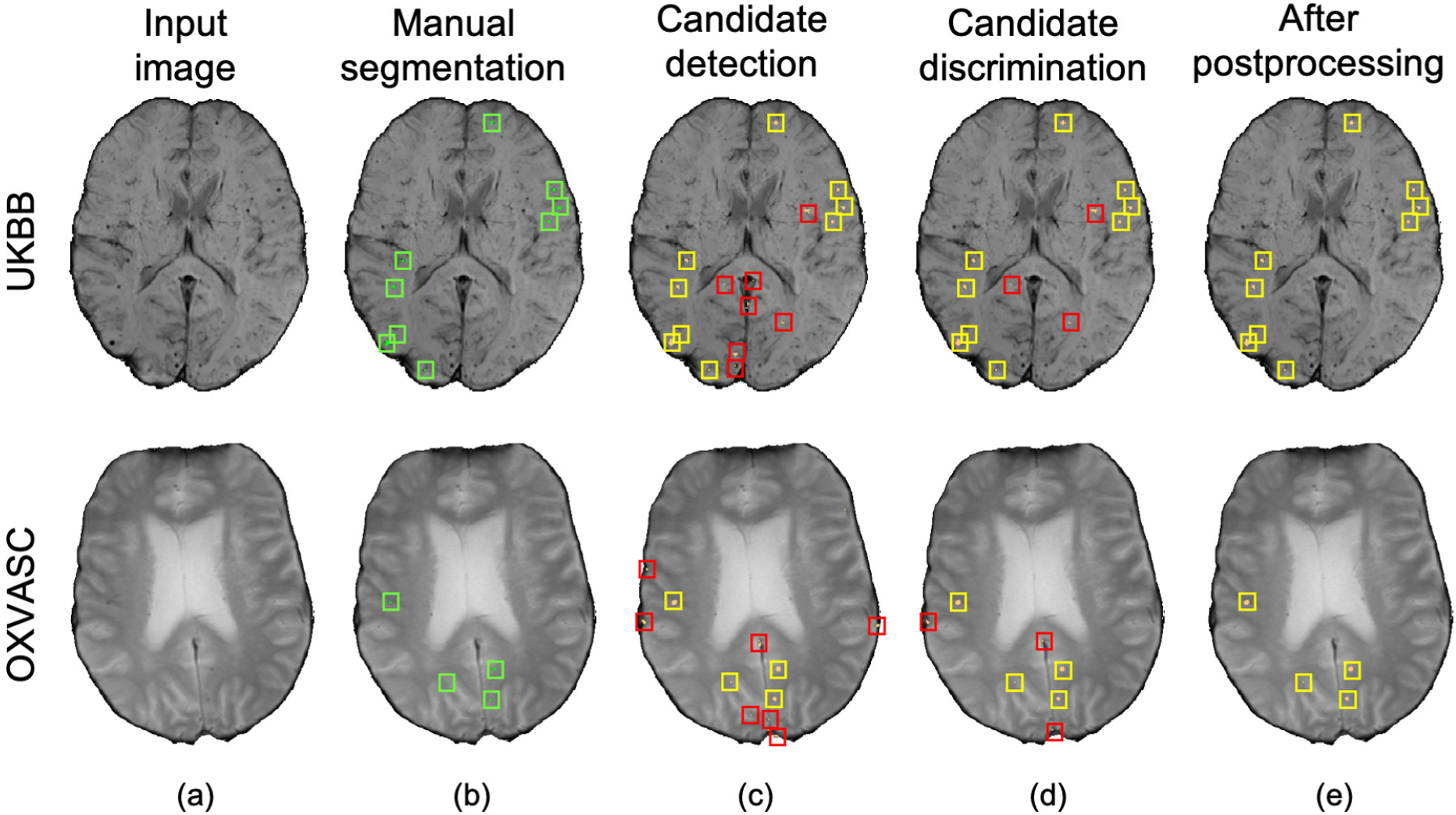
Sample cross-validation results on the UKBB (top panel) and the OXVASC (bottom panel) datasets. (a) Input image and (b) manual segmentations shown along with results at (c) CMB initial candidate detection, (d) candidate discrimination and (e) after postprocessing steps. True positive and false positive candidates are shown in yellow and red boxes respectively for each step.

### 5.3. Evaluation of the generalisability of the proposed method

Table 4 reports the performance metrics of the proposed method, when trained on the UKBB dataset and applied on the same dataset but different modality (UKBB QSM data), different datasets but same modality (SWI from the TICH2 and SHK datasets) and different dataset and modality (T2*-GRE from the OXVASC dataset). We used *Th*_*Cdet*_ and *Th*_*Cdisc*_ values of 0.3 (determined from the cross-validation on the UKBB dataset) on the probability maps at the candidate detection step and on the patch-level probabilities at the discrimination step. Figure 9 shows sample results of the method, when applied on various datasets at various steps of CMB detection.

**Table 4:** Evaluation of the generalisability of the proposed method - trained on the UKBB SWI data and evaluated on the UKBB QSM, TICH2, SHK and OXVASC datasets: performance metrics at candidate detection, discrimination and post-processing steps. Cl. TPR and Cl. prec indicate cluster-wise TPR and cluster-wise precision respectively. C. det - candidate detection, C. disc - candidate discrimination.

| Datasets | Steps | Cl. TPR | FPavg | Cl. prec |
| --- | --- | --- | --- | --- |
| **UKBB (QSM)** | C. det. | 0.99 | 138.0 | 0.02 |
|  | C. disc. | 0.91 | 40.3 | 0.04 |
|  | Postproc. | 0.90 | 1.8 | 0.44 |
| **TICH2 (SWI)** | C. det. | 0.88 | 289.3 | 0.02 |
|  | C. disc. | 0.83 | 42.8 | 0.10 |
|  | Postproc. | 0.82 | 3.1 | 0.62 |
| **SHK (SWI)** | C. det. | 0.98 | 254.7 | 0.01 |
|  | C. disc. | 0.94 | 43.6 | 0.09 |
|  | Postproc. | 0.87 | 0.5 | 0.89 |
| **OXVASC (T2\*-GRE)** | C. det. | 0.88 | 147.1 | 0.03 |
|  | C. disc. | 0.85 | 53.7 | 0.07 |
|  | Postproc. | 0.81 | 2.0 | 0.71 |

**Figure 9:**
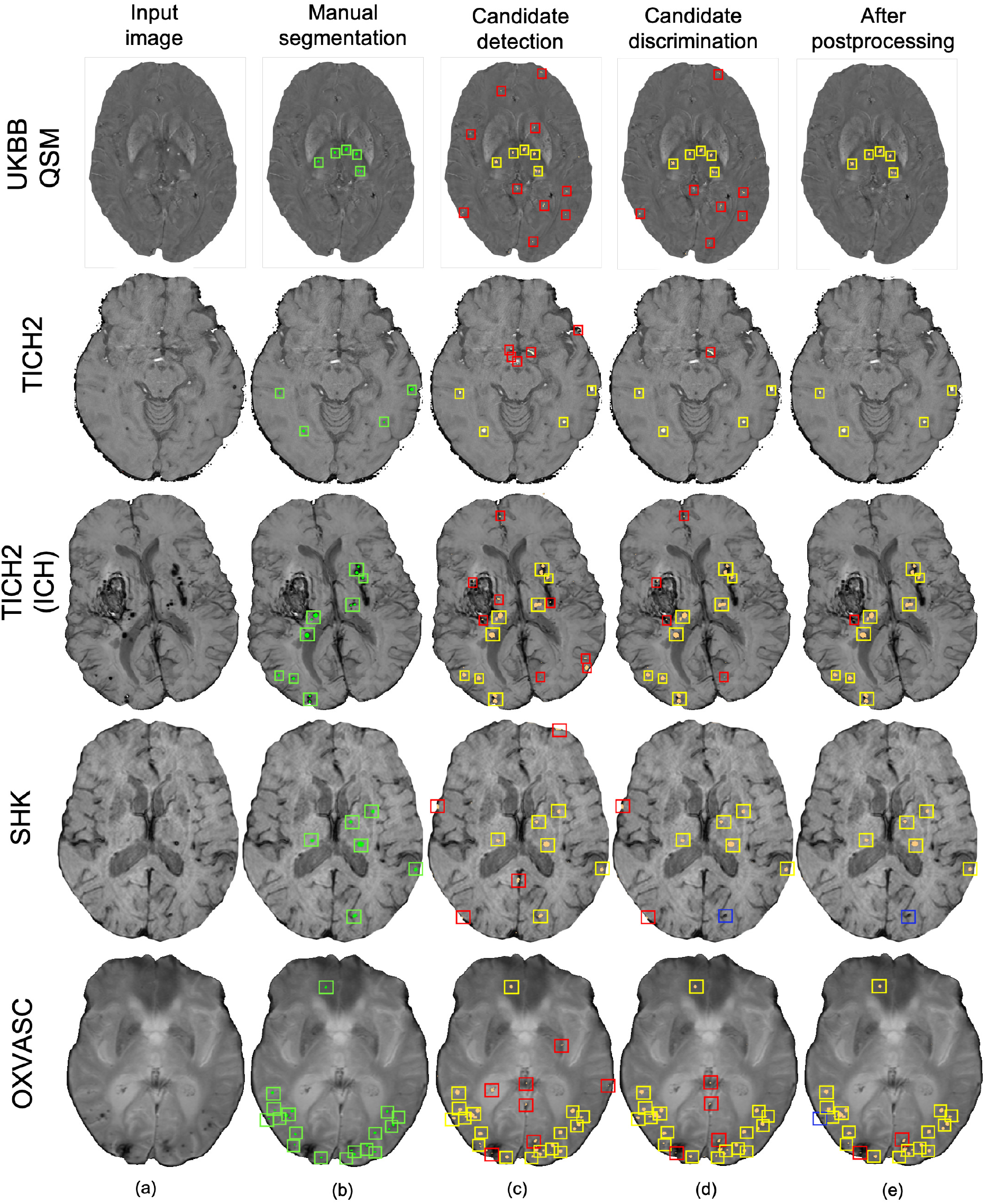
Sample results of the UKBB-trained method on the UKBB QSM, TICH2 (without and with ICH), SHK and OXVASC datasets (from top to bottom panels). (a) Input image and (b) manual segmentations shown along with results at (c) CMB initial candidate detection, (d) candidate discrimination and (e) after postprocessing steps. True positive, false positive and false negative candidates are shown in yellow, red and blue boxes respectively for each step.

Out of all datasets, the method achieved the highest cluster-wise TPR on the QSM dataset. On this dataset, the results were on par with the cross-validation results on the UKBB SWI data (with slight decrease in the cluster-wise TPR and precision on QSM data). We obtained FPavg values of 1.8 FPs/subject - the FPs candidates were mainly due to minor susceptibility changes in the tissue and penetrating small blood vessels.

The method gave a cluster-wise TPR of 0.82 on the TICH2 dataset, despite the presence of ICH lesions (third row in figure 9) in all subjects. The method provided the highest FPavg values in the initial candidate detection step (FPavg = 289.3 FPs/subject), possibly due to ICH edges and texture. However, the candidate discrimination step reduced the number of FPs and lowered the FPavg value to 42.8 FP/subject, which is comparable with other datasets. Even then, we obtained the highest FPavg after post-processing on this dataset with a cluster-wise precision of 0.62. Susceptibility artefacts at tissue interfaces and sulci were mainly detected as FPs in this dataset.

On the SHK dataset, while the first two steps (CMB detection and discrimination) provided consistently good cluster-wise TPR values (*>*0.90), the TPR value decreased at the post-processing step. Even then, on this dataset our method provided the lowest FPavg (0.5 FPs/subject) and the highest cluster-wise precision among all the datasets. The FPs in the candidate detection step were mostly regions of intersections of blood vessels closer to sulci, especially the central sulcus.

On the OXVASC dataset, the method achieved the lowest cluster-wise TPR of 0.81. The high number of false negatives in this dataset, as suggested by the lower cluster-wise TPR, could be due to the reduced contrast between CMBs and normal brain tissue, unlike the SWI data used for training. Also, occasionally true CMB candidates quite close to the skull were removed in the post-processing step, having been mistaken as sulci.

### 5.4. Comparison with existing methods

Table 5 provides a comparison of the proposed CMB detection method with existing fully automated methods. From the table, generally deep-learning-based methods performed better compared to conventional machine learning methods. Also, the methods using multiple modalities or using phase information in addition to SWI (Liu et al., 2019b; Ghafaryasl et al., 2012; Al-Masni et al., 2020; Rashid et al., 2021) showed better results. In fact, Al-Masni et al. (2020) showed that using phase in addition to SWI images improves the cluster-wise TPR by 5.6% (with only SWI: 91.6% and with SWI and phase: 97.2%) and Rashid et al. (2021) achieved the best CMB detection performance by using T2-weighted, SWI and QSM modalities. However, our proposed method uses a single modality (SWI or T2*-GRE), along with the FRST images (obtained from the input modality itself) and gives comparable results to state-of-the-art methods such as Al-Masni et al. (2020) and Liu et al. (2019b), and with lower FPavg compared to Dou et al. (2016). Also, our precision values on both UKBB and OXVASC datasets are better than existing methods including Dou et al. (2016); Bian et al. (2013); Fazlollahi et al. (2014, 2015). We have reported an additional comparison of patch-level methods in the supplementary material. While the methods using patch-wise evaluation (Chen et al., 2018; Lu et al., 2021b) provided good patch-level sensitivity and accuracies (shown in the supplementary material), we cannot compare the performance of our method with those, since they used preselected CMB patches (from manually annotated CMB voxels) and comparable numbers of non-CMB patches as inputs. Also the input CMB patches occasionally contained multiple CMBs, which makes the comparison with cluster-wise metrics highly difficult.

**Table 5:** Comparison of the performance of the proposed CMB detection method with existing conventional machine learning (ML) and deep learning (DL) methods. Cl.TPR - cluster-wise TPR, FPavg - Average false positives per image/subject, Cl.Prec - cluster-wise precision, FPavg_*CMB*_ - FPavg for CMB subjects, FPavg_*nCMB*_ - FPavg for non-CMB subjects, FPavg_*D*_ - FPavg for ‘*definite*’ CMB subjects, FPavg_*P* +*D*_ - FPavg for ‘*definite and possible*’ CMB subjects.

| Methods | Datasets |  | Performance |  |  |
| --- | --- | --- | --- | --- | --- |
|  | Sequence(s)<br>(# test subjects) | Total<br>#<br>CMBs | CMB<br>screening step | CMB<br>discrimination step | FP reduction<br>step |
| **ML methods** |  |  |  |  |  |
| Bian et al. (2013) | SWI (10) | 304 | FPavg - 287.7 |  | Cl.TPR - 86.5%,<br>FPavg - 44.9 |
| Fazlollahi et al. (2014) | SWI (41) | 103 | Cl.TPR - 98%,<br>FPavg - 695 | Cl.TPR - 92%,<br>FPavg<sub>CMB</sub> - 6.7<br>FPavg<sub>nCMB</sub> - 16.8 |  |
| Fazlollahi et al. (2015) | SWI (66) | 231 | Cl.TPR - 97%,<br>FPavg<sub>D</sub> - 669,<br>FPavg<sub>P+D</sub> - 1373 | Cl.TPR - 87%,<br>FPavg<sub>D</sub> - 10.28,<br>FPavg<sub>P+D</sub> - 27.8 |  |
| Ghafaryasl et al. (2012) | T2*-GRE + PD (81) | 183 | Cl.TPR - 98%,<br>FPavg - 705 | Cl.TPR - 92%,<br>FPavg - 19 | Cl.TPR - 91%,<br>FPavg - 4.1 |
| Dou et al. (2015) | SWI (19) | 161 | Cl.TPR - 100%,<br>FPavg - 807 | Cl.TPR - 94%,<br>FPavg - 190 | Cl.TPR - 80%,<br>FPavg - 7.7,<br>Cl.Prec - 49% |
| Chesebro et al. (2021) | T2*-GRE, SWI (78) | 64 |  |  | Cl.TPR - 95%,<br>FPavg - 9.7<br>(SWI), 17.1<br>(T2*-GRE),<br>Cl.prec - 11%<br>(SWI), 7%<br>(T2*-GRE) |
| **DL methods** |  |  |  |  |  |
| Chen et al. (2015) | SWI (5) | 55 | Cl.TPR - 96%,<br>FPavg > 800 | Cl.TPR - 89%,<br>FPavg - 6.4,<br>Cl.Prec - 56% |  |
| Dou et al. (2016) | SWI (50) | 117 | Cl.TPR - 98%,<br>FPavg - 282 | Cl.TPR - 93%, FPavg - 2.74, Cl.Prec - 44% |  |
| Liu et al. (2019b) | Phase + SWI (41) | 168 | Cl.TPR - 99%,<br>FPavg - 276.8 |  | Cl.TPR - 96%,<br>FPavg - 1.8 |
| Al-Masni et al. (2020) (5-fold cross-validation) | Phase + SWI (72) | 188 | Cl.TPR - 93.62%, FPavg - 52.18 | Cl.TPR - 94.32% of candidates detected in step 1, FPavg - 1.4, Cl.Prec - 61.9% |  |
| Rashid et al. (2021) (Leave-one-out cross-validation) | QSM + SWI + T2w (24) | ≈172 | Cl.TPR - 88%,<br>Cl.Prec - 40% |  |  |
| **Proposed method** (5-fold cross-validation) |  |  |  |  |  |
|  | UKBB - SWI (78) | 186 | Cl.TPR - 97%,<br>FPavg - 175.4 | Cl.TPR - 95%,<br>FPavg - 24.2 | Cl.TPR - 93%,<br>FPavg - 1.5,<br>Cl.Prec - 59% |
|  | OXVASC - T2*-GRE (74) | 366 | Cl.TPR - 93%,<br>FPavg - 195.7 | Cl.TPR - 91%,<br>FPavg - 10.1 | Cl.TPR - 90%,<br>FPavg - 0.9,<br>Cl.Prec - 84% |

## 6. Discussion and conclusions

In this work, we proposed a fully automated, deep-learning-based, 2-step method for accurate lesion-level detection of CMBs on various datasets, irrespective of variations in population-level, scanner and acquisition characteristics. Our method uses a single input modality and the radial symmetry property of CMBs for detection of CMB candidates with high sensitivity in the initial candidate detection step. For the candidate discrimi-nation step, we trained a student classification network with the knowledge distilled from a multi-tasking teacher network for accurate classification of CMB candidates from non-CMB candidates. Our ablation study results show that the candidate discrimination and post-processing steps drastically reduce the number of FPs, and that the use of the KD framework improves cluster-wise TPR values at the discrimination step. Our method achieved cluster-wise TPR values *>*90% with FPavg of *<*1.6 FPs/subject during initial cross-validation on the UKBB and OXVASC datasets consisting of SWI and T2*-GRE images respectively. On training the models on the UKBB dataset and applying them on different datasets with different demographic and scanner-related variations, the method showed a good generalisability across datasets, providing cluster-wise TPR values *>*80% on all datasets.

The initial vessel removal preprocessing step helped in reducing the number of FPs, since blood vessels (especially the small ones closer to sulci) are one of the common mimics of CMBs. One of the main challenges in the vessel removal step is the potential removal of true CMBs that are very close to vessels. Hence, we removed only linear segments with low uniform width in this step. Therefore, this step removed the vessels and sulci that were more prominent and could lead to obvious FPs. This was especially effective for the SHK dataset, where the blood vessels and sulci had higher contrast and were distinctly different from CMBs (figure 1b). While removing the linear, elongated structures from the images, we also aimed to leverage the radial symmetry property of CMBs. Towards that aim, using FRST maps, obtained from the input modality, as an additional input channel helped the candidate detection model in learning contextual features, leading to the detection of more true positive CMBs. This was particularly useful, given the lack of location prior for the CMB distribution in the brain.

The main objective of the candidate detection step is to detect as many true positive candidates as possible, with a trade-off of high FPavg, because any of the CMB candidates missed in this stage cannot be recovered in the subsequent steps. From the ablation study, given high FPavg in the candidate detection step, the student model trained using KD reduced FPavg approximately by a factor of 4 with a smaller decrease in cluster-wise TPR, when compared to the classification model trained without KD. The advantage of the teacher model in the proposed discrimination step was 2-fold: (1) its ability to learn the contextual features that are salient for both voxel-wise CMB detection and patch-level CMB/non-CMB classification and (2) the use of a multi-tasking framework that provides a regularisation effect on the tasks, reducing the chance of over-fitting and false classifications. We observed that the multi-tasking framework, together with the upweighting of the CMB classes in the loss function, reduced the effect of class imbalance between CMB and non-CMB patches (note that the model trained without KD is slightly biased towards the non-CMB class, evident from the lower cluster-wise TPR and FPavg values in table 2). Therefore, using the teacher model to train the student model further enhanced the capability of the network in differentiating CMB and non-CMB patches (as shown by feature embeddings in t-SNE plots in figure 6c, d). Additionally, we also provided the input patches centered on detected initial CMB candidates to the student model. This, in addition to the distilled knowledge from the teacher, enabled the model to focus on the pattern at the centre of the patches for accurate classification of CMB patches. This was especially useful to remove the fragments of blood vessels (e.g. intersections and branching points) missed in the vessel removal step as seen in figures 8d and 9d. Regarding the parameters used in KD, using a higher temperature (*τ*) results in softened softmax values between classes and has been shown to typically provide the knowledge (also known as *dark knowledge*) for training a generalisable student network (Hinton et al., 2015). However, given the similarities in the characteristics of CMBs and mimics, having a very high *τ* values could lead to misclassifications. Our main aim was to achieve a good hard prediction to differentiate the CMB class from the non-CMB class, while at the same time to transfer the knowledge from the teacher model to the student model. Hence we empirically chose an optimal *τ* value of 4 (that provided smoother softmax values without affecting the CMB/non-CMB prediction) based on manual tuning. Further removal of FPs in the post-processing resulted in the drastic improvement in the cluster-wise precision. Noise reduction or smoothing during preprocessing might lead to a loss of CMBs (even for data augmentation, very small *σ* values were chosen carefully). Therefore, small intensity and texture variations (mainly in the sub-cortical and lobar regions) led to detection of FPs, which were removed in the post-processing step.

As for the cross-validation results, the method achieved the highest cluster-wise TPR values on SWI images (from the UKBB dataset), while providing the lowest FPavg and the highest cluster-wise precision on T2*-GRE images (from the OXVASC dataset). This could be due to fact that CMBs appear with a higher contrast on SWI compared to T2*-GRE images due to the blooming effect. This also affects most of the CMB mimics as well, increasing their contrast on SWI, leading to high cluster-wise TPR but also high FPavg. Also, T2*-GRE images had a smoother texture when compared to SWI (figure 8), resulting in less noisy FRST maps, hence leading to the improved performance metrics at the candidate discrimination step in the OXVASC dataset. However, the FPavg value at the initial candidate detection step was higher for the OXVASC dataset due to the lower voxel resolution in the z-direction (5mm) in the OXVASC data, leading to partial volume artefacts and making it highly difficult to differentiate between small sulci closer to the skull and CMBs.

On evaluating the generalisability of our method on various datasets, our method trained on the SWI data from the UKBB dataset showed good generalisability on QSM images from the same dataset, with comparable performance to the cross-validation results on the SWI data. Regarding the performance after individual steps, in the initial candidate detection step, the method provided the highest cluster-wise TPR values with the lowest FPavg values on the QSM data (even lower than with UKBB SWI data) since QSM shows better separation of diamagnetic mimics from CMBs. However, due to local tissue susceptibility variations (which is quite different from the SWI training data), the FPavg in the candidate discrimination step was higher that it was when using the SWI data. Finally, the post-processing step effectively removed the stray voxels due to noisy susceptibility variations (that were extremely small and hence were below the 5-voxel threshold) and reduced the FPavg value to 1.8 FPs/subject. It is worth noting that, since the same subjects were used for training (SWI data for training and QSM data for testing), the results are likely to be biased. That is, the model could have learnt the overall locations of CMBs for the training subjects, rather than the modality-invariant features. However, we believe that the use of patches, rather than whole slices or volumes, at both steps would reduce the chance of biased assessment.

For the datasets consisting of the same modality as that of the training data (SWI) but from different populations, the method was affected by the presence of additional pathological signs (e.g. ICH in the TICH2 dataset). In the TICH2 dataset, the noisy texture of the haemorrhage regions and their edges led to the highest FPavg value in the initial candidate detection step. In terms of FPs, we found that additional pathological signs, that were not the part of training, affected the method more than the modalities. For instance, among the OXVASC (different modality from the training SWI data) and TICH2 datasets (same modality), even though both are pathological datasets, the greater prevalence of confounding ‘CMB-like’ signs in TICH2 resulted in higher FPavg in the TICH2 dataset. Among all the datasets we used, the SHK dataset had high contrast, low noise and a better than average resolution making vessels and sulci easy to remove in this dataset. Moreover, this dataset has same modality as that of the training data, and hence both candidate detection and discrimination step models performed well (and cluster-wise TPR values comparable even with that on the UKBB SWI data). However, during the post-processing step, a few CMBs near the sulci, closer to the skull were misclassified as FPs resulting in lower cluster-wise TPR. The OXVASC dataset was quite different from the training SWI data and from other datasets, since it shows lower contrast between CMB and background as shown in figure 9. Hence, providing FRST as the second input channel was particularly useful for this dataset, since the FRST relies more on the radial symmetry nature of CMBs at different radii (we used 2, 3, 4 and 6 as specified in section 2.2) rather than its intensity contrast with respect to the background. Hence, on the OXVASC dataset the FRST maps had the same contrast as that of other modalities (as seen in figure 2a) aiding in the detection of subtle CMBs. Since the estimation of FRST maps does not require any additional modality other than the input modality, our method effectively uses a single image modality and provided results comparable to existing methods that use multiple modalities (Ghafaryasl et al., 2012; Al-Masni et al., 2020).

Concluding, we proposed a fully automated method using deep learning for CMB candidate detection, candidate discrimination with a knowledge distillation framework, followed by post-processing filtering using structural and anatomical properties. Our method achieved cluster-wise TPR values of *>*90% with FPavg *<*1.6 FPs/subject on T2*-GRE and SWI modalities, on par with the state-of-the-art, and gave better precision than existing methods. When the models were trained on SWI data and applied on QSM images from the same dataset, the method achieved a cluster-wise TPR *≈*90%. On applying the trained method to other datasets consisting of data from different populations and acquired using different scanners and protocols, our method gave a cluster-wise TPR *>*81%, despite the presence of other major pathologies. The python implementation of the codes for CMB candidate detection and discrimination steps are currently available in https://github.com/v-sundaresan/microbleed-detection. One of the future directions of this research would be to improve the generalisability of the proposed method using various domain adaptation techniques, to overcome the effect of scanner- and population-related variations. Another clinically focussed avenue of this research could be to develop automated algorithms to rate the CMBs based on their size and distribution, which would be useful in studying their clinical impact.

## Data Availability

Requests for data from the OXVASC Study will be considered by P.M.R. in line with data protection laws. The TICH-2 MRI sub-study data can be shared with bona fide researchers and research groups on written request to the sub-study PI Prof Rob Dineen. Proposals will be assessed by the PI (with advice from the TICH-2 trial Steering Committee if required) and a Data Transfer Agreement will established before any data are shared. The UK Biobank and Hong Kong (HK) datasets are available to researchers through an open applications via https://www.ukbiobank.ac.uk/register-apply/ and http://www.cse.cuhk.edu.hk/~qdou/cmb-3dcnn/cmb-3dcnn.html respectively

## Acknowledgements

This research was funded in part by the Wellcome Trust [203139/Z/16/Z]. For the purpose of open access, the author has applied a CC-BY public copyright licence to any Author Accepted Manuscript version arising from this submission. This work was also supported by the Engineering and Physical Sciences Research Council (EPSRC), Medical Research Council (MRC) [grant number EP/L016052/1], NIHR Nottingham Biomedical Research Centre and Wellcome Centre for Integrative Neuroimaging, which has core funding from the Wellcome Trust [203139/Z/16/Z]. The computational aspects of this research were funded from National Institute for Health Research (NIHR) Oxford BRC with additional support from the Wellcome Trust Core Award Grant Number 203141/Z/16/Z. The Oxford Vascular Study is funded by the National Institute for Health Research (NIHR) Oxford Biomedical Research Centre (BRC), Wellcome Trust, Wolfson Foundation, the British Heart Foundation and the European Unions Horizon 2020 programme (grant 666881, SVDs@target). The TICH-2 MRI sub-study was funded by a grant from British Heart Foundation (grant reference PG/14/96/31262) and the TICH-2 trial was funded by a grant from the NIHR Health Technology Assessment programme (project code 11 129 109). The views expressed are those of the author(s) and not necessarily those of the NHS, the NIHR or the Department of Health. VS is supported by the Wellcome Centre for Integrative Neuroimaging [203139/Z/16/Z]. CA was supported by NIHR Nottingham Biomedical Research Centre and is now supported by Wellcome Trust Collaborative Award [215573/Z/19/Z]. GZ is supported by the Italian Ministry of Education (MIUR) and by a grant Dipartimenti di eccellenza 2018-2022, MIUR, Italy, to the Department of Biomedical, Metabolic and Neural Sciences, University of Modena and Reggio Emilia. PMR is in receipt of a NIHR Senior Investigator award. KM and BT are funded by a Senior Research Fellowship from the Wellcome Trust (202788/Z/16/Z). MJ is supported by the NIHR Oxford Biomedical Research Centre (BRC), and this research was funded by the Wellcome Trust (215573/Z/19/Z). LG is supported by the National Institute for Health Research (NIHR) Oxford Health Biomedical Research Centre (BRC).

The UKBB data used in this work was obtained from UK Biobank under Data Access Applications (8107, 43822). We are grateful to the UK Biobank for making the resource data available, and are extremely grateful to all UK Biobank study participants, who generously donated their time to make this resource possible. We acknowledge all the OXVASC study participants. For the OXVASC dataset, we acknowledge the use of the facilities of the Acute Vascular Imaging Centre, Oxford. We also thank Dr. Chiara Vincenzi and Dr. Francesco Carletti for their help on generating the manual masks used in our experiments.

MJ receives royalties from licensing of FSL to non-academic, commercial parties. The authors report no potential conflicts of interest.

## References

Al-Masni, M.A., Kim, W.R., Kim, E.Y., Noh, Y., Kim, D.H., 2020. A two cascaded network integrating regional-based yolo and 3d-cnn for cerebral microbleeds detection, in: 2020 42nd Annual International Conference of the IEEE Engineering in Medicine & Biology Society (EMBC), IEEE. pp. 1055–1058.

Ba, L.J., Caruana, R., 2013. Do deep nets really need to be deep? arXiv preprint 1312.6184.

Barnes, S.R., Haacke, E.M., Ayaz, M., Boikov, A.S., Kirsch, W., Kido, D., 2011. Semiautomated detection of cerebral microbleeds in magnetic resonance images. Magnetic resonance imaging 29, 844–852.

Bian, W., Hess, C.P., Chang, S.M., Nelson, S.J., Lupo, J.M., 2013. Computer-aided detection of radiation-induced cerebral microbleeds on susceptibility-weighted MR images. NeuroImage: clinical 2, 282–290.

Bucilu, C., Caruana, R., Niculescu-Mizil, A., 2006. Model compression, in: Proceedings of the 12th ACM SIGKDD international conference on Knowledge discovery and data mining, pp. 535–541.

Charidimou, A., Werring, D.J., 2011. Cerebral microbleeds: detection, mechanisms and clinical challenges. Future Neurology 6, 587–611.

Chen, G., Choi, W., Yu, X., Han, T., Chandraker, M., 2017. Learning efficient object detection models with knowledge distillation, in: Proceedings of the 31st International Conference on Neural Information Processing Systems, pp. 742–751.

Chen, H., Yu, L., Dou, Q., Shi, L., Mok, V.C., Heng, P.A., 2015. Automatic detection of cerebral microbleeds via deep learning based 3D feature representation, in: 2015 IEEE 12th international symposium on biomedical imaging (ISBI), IEEE. pp. 764–767.

Chen, Y., Villanueva-Meyer, J.E., Morrison, M.A., Lupo, J.M., 2018. Toward automatic detection of radiation-induced cerebral microbleeds using a 3d deep residual network. Journal of digital imaging, 1–7.

Chesebro, A.G., Amarante, E., Lao, P.J., Meier, I.B., Mayeux, R., Brickman, A.M., 2021. Automated detection of cerebral microbleeds on t2*-weighted mri. Scientific reports 11, 1–13.

Cordonnier, C., Wardlaw, J., Al-Shahi Salman, R., 2007. Spontaneous brain microbleeds: systematic review, subgroup analyses and standards for study design and reporting. Brain 130, 1988–2003. URL: https://doi.org/10.1093/brain/awl387, doi:10.1093/brain/awl387.

De Bresser, J., Brundel, M., Conijn, M., Van Dillen, J., Geerlings, M., Viergever, M., Luijten, P., Biessels, G., 2013. Visual cerebral microbleed detection on 7T MR imaging: reliability and effects of image processing. American Journal of Neuroradiology 34, E61–E64.

Dineen, R.A., Pszczolkowski, S., Flaherty, K., Law, Z.K., Morgan, P.S., Roberts, I., Werring, D.J., Salman, R.A.S., England, T., Bath, P.M., et al., 2018. Does tranexamic acid lead to changes in MRI measures of brain tissue health in patients with spontaneous intracerebral haemorrhage? protocol for a MRI substudy nested within the double-blind randomised controlled TICH-2 trial. BMJ open 8, e019930.

Ding, Q., Wu, S., Sun, H., Guo, J., Xia, S.T., 2019. Adaptive regularization of labels. arXiv preprint 1908.05474.

Dou, Q., Chen, H., Yu, L., Shi, L., Wang, D., Mok, V.C., Heng, P.A., 2015. Automatic cerebral microbleeds detection from MR images via independent subspace analysis based hierarchical features, in: 2015 37th annual international conference of the IEEE engineering in medicine and biology society (EMBC), IEEE. pp. 7933–7936.

Dou, Q., Chen, H., Yu, L., Zhao, L., Qin, J., Wang, D., Mok, V.C., Shi, L., Heng, P.A., 2016. Automatic detection of cerebral microbleeds from MR images via 3d convolutional neural networks. IEEE transactions on medical imaging 35, 1182–1195.

Fazlollahi, A., Meriaudeau, F., Giancardo, L., Villemagne, V.L., Rowe, C.C., Yates, P., Salvado, O., Bourgeat, P., Group, A.R., et al., 2015. Computer-aided detection of cerebral microbleeds in susceptibility-weighted imaging. Computerized Medical Imaging And Graphics 46, 269–276.

Fazlollahi, A., Meriaudeau, F., Villemagne, V.L., Rowe, C.C., Yates, P., Salvado, O., Bourgeat, P., 2014. Efficient machine learning framework for computer-aided detection of cerebral microbleeds using the radon transform, in: 2014 IEEE 11th international symposium on biomedical imaging (ISBI), IEEE. pp. 113–116.

Förstner, W., 1994. A framework for low level feature extraction, in: European Conference on Computer Vision, Springer. pp. 383–394.

Frangi, A.F., Niessen, W.J., Vincken, K.L., Viergever, M.A., 1998. Multiscale vessel enhancement filtering, in: International conference on medical image computing and computer-assisted intervention, Springer. pp. 130–137.

Ganin, Y., Ustinova, E., Ajakan, H., Germain, P., Larochelle, H., Laviolette, F., Marchand, M., Lempitsky, V., 2016. Domain-adversarial training of neural networks. The Journal of Machine Learning Research 17, 2096–2030.

Ghafaryasl, B., van der Lijn, F., Poels, M., Vrooman, H., Ikram, M.A., Niessen, W.J., van der Lugt, A., Vernooij, M., de Bruijne, M., 2012. A computer aided detection system for cerebral microbleeds in brain MRI, in: 2012 9th IEEE international symposium on biomedical imaging (ISBI), IEEE. pp. 138–141.

Greenberg, S.M., Vernooij, M.W., Cordonnier, C., Viswanathan, A., Salman, R.A.S., Warach, S., Launer, L.J., Van Buchem, M.A., Breteler, M.M., Group, M.S., et al., 2009. Cerebral microbleeds: a guide to detection and interpretation. The Lancet Neurology 8, 165–174.

Gregoire, S., Chaudhary, U., Brown, M., Yousry, T., Kallis, C., Jäger, H., Werring, D., 2009. The Microbleed Anatomical Rating Scale (MARS): reliability of a tool to map brain microbleeds. Neurology 73, 1759–1766.

Guo, Q., Wang, X., Wu, Y., Yu, Z., Liang, D., Hu, X., Luo, P., 2020. Online knowledge distillation via collaborative learning, in: Proceedings of the IEEE/CVF Conference on Computer Vision and Pattern Recognition, pp. 11020–11029.

Haacke, E.M., Xu, Y., Cheng, Y.C.N., Reichenbach, J.R., 2004. Susceptibility weighted imaging (swi). Magnetic Resonance in Medicine: An Official Journal of the International Society for Magnetic Resonance in Medicine 52, 612–618.

He, K., Zhang, X., Ren, S., Sun, J., 2016. Deep residual learning for image recognition, in: Proceedings of the IEEE conference on computer vision and pattern recognition, pp. 770–778.

van den Heuvel, T., Van Der Eerden, A., Manniesing, R., Ghafoorian, M., Tan, T., Andriessen, T., Vyvere, T.V., Van den Hauwe, L., ter Haar Romeny, B., Goraj, B., et al., 2016. Automated detection of cerebral microbleeds in patients with traumatic brain injury. NeuroImage: Clinical 12, 241–251.

Hinton, G., Vinyals, O., Dean, J., 2015. Distilling the knowledge in a neural network. arXiv preprint 1503.02531.

Hong, J., Wang, S.H., Cheng, H., Liu, J., 2020. Classification of cerebral microbleeds based on fully-optimized convolutional neural network. Multimedia Tools and Applications 79, 15151–15169.

Hu, M., Maillard, M., Zhang, Y., Ciceri, T., La Barbera, G., Bloch, I., Gori, P., 2020. Knowledge distillation from multi-modal to mono-modal segmentation networks, in: International Conference on Medical Image Computing and Computer-Assisted Intervention, Springer. pp. 772–781.

Jin, X., Peng, B., Wu, Y., Liu, Y., Liu, J., Liang, D., Yan, J., Hu, X., 2019. Knowledge distillation via route constrained optimization, in: Proceedings of the IEEE/CVF International Conference on Computer Vision, pp. 1345–1354.

Kim, S.W., Kim, H.E., 2017. Transferring knowledge to smaller network with class-distance loss.

Kingma, D.P., Ba, J., 2014. Adam: A method for stochastic optimization. arXiv preprint 1412.6980.

Kuijf, H.J., de Bresser, J., Geerlings, M.I., Conijn, M.M., Viergever, M.A., Biessels, G.J., Vincken, K.L., 2012. Efficient detection of cerebral microbleeds on 7.0 T MR images using the radial symmetry transform. NeuroImage 59, 2266–2273.

Kuijf, H.J., Brundel, M., de Bresser, J., van Veluw, S.J., Heringa, S.M., Viergever, M.A., Biessels, G.J., Vincken, K.L., 2013. Semi-automated detection of cerebral microbleeds on 3.0 T MR images. PLoS One 8, e66610.

Lachinov, D., Shipunova, E., Turlapov, V., 2019. Knowledge distillation for brain tumor segmentation, in: International MICCAI Brainlesion Workshop, Springer. pp. 324–332.

Lan, X., Zhu, X., Gong, S., 2018. Knowledge distillation by on-the-fly native ensemble. arXiv preprint 1806.04606.

Li, Y., Yang, J., Song, Y., Cao, L., Luo, J., Li, L.J., 2017. Learning from noisy labels with distillation, in: Proceedings of the IEEE International Conference on Computer Vision, pp. 1910–1918.

Liu, C., Li, W., Tong, K.A., Yeom, K.W., Kuzminski, S., 2015. Susceptibility-weighted imaging and quantitative susceptibility mapping in the brain. Journal of magnetic resonance imaging 42, 23–41.

Liu, L., Wang, H., Lin, J., Socher, R., Xiong, C., 2019a. Mkd: a multi-task knowledge distillation approach for pretrained language models. arXiv preprint 1911.03588.

Liu, S., Utriainen, D., Chai, C., Chen, Y., Wang, L., Sethi, S.K., Xia, S., Haacke, E.M., 2019b. Cerebral microbleed detection using susceptibility weighted imaging and deep learning. NeuroImage 198, 271–282.

Loy, G., Zelinsky, A., 2002. A fast radial symmetry transform for detecting points of interest, in: European Conference on Computer Vision, Springer. pp. 358–368.

Lu, D., Liu, J., MacKinnon, A.D., Tozer, D.J., Markus, H.S., 2021a. Prevalence and risk factors of cerebral microbleeds: Analysis from the uk biobank. Neurology 97, e1493–e1502.

Lu, S.Y., Nayak, D.R., Wang, S.H., Zhang, Y.D., 2021b. A cerebral microbleed diagnosis method via featurenet and ensembled randomized neural networks. Applied Soft Computing, 107567.

Morrison, M.A., Payabvash, S., Chen, Y., Avadiappan, S., Shah, M., Zou, X., Hess, C.P., Lupo, J.M., 2018. A user-guided tool for semi-automated cerebral microbleed detection and volume segmentation: Evaluating vascular injury and data labelling for machine learning. NeuroImage: Clinical 20, 498–505.

Müller, R., Kornblith, S., Hinton, G., 2019. When does label smoothing help? arXiv preprint 1906.02629.

Nandigam, R., Viswanathan, A., Delgado, P., Skehan, M., Smith, E., Rosand, J., Greenberg, S., Dickerson, B., 2009. MR imaging detection of cerebral microbleeds: Effect of susceptibility-weighted imaging, section thickness, and field strength. American Journal of Neuroradiology 30, 338–343. URL: http://www.ajnr.org/content/30/2/338, doi:10.3174/ajnr.A1355, arXiv:http://www.ajnr.org/content/30/2/338.full.pdf.

Pan, Z.W., Xiang, D.H., Xiao, Q.W., Zhou, D.X., 2008. Parzen windows for multi-class classification. Journal of complexity 24, 606–618.

Rashid, T., Abdulkadir, A., Nasrallah, I.M., Ware, J.B., Liu, H., Spincemaille, P., Romero, J.R., Bryan, R.N., Heckbert, S.R., Habes, M., 2021. Deepmir: a deep neural network for differential detection of cerebral microbleeds and iron deposits in mri. Scientific reports 11, 1–14.

Redmon, J., Farhadi, A., 2017. Yolo9000: better, faster, stronger, in: Proceedings of the IEEE conference on computer vision and pattern recognition, pp. 7263–7271.

Romero, A., Ballas, N., Kahou, S.E., Chassang, A., Gatta, C., Bengio, Y., 2014. Fitnets: Hints for thin deep nets. arXiv preprint 1412.6550.

Rothwell, P., Coull, A., Giles, M., Howard, S., Silver, L., Bull, L., Gutnikov, S., Edwards, P., Mant, D., Sackley, C., et al., 2004. Change in stroke incidence, mortality, case-fatality, severity, and risk factors in Oxfordshire, UK from 1981 to 2004 (Oxford Vascular Study). The Lancet 363, 1925–1933.

Sarfraz, F., Arani, E., Zonooz, B., 2019. Noisy collaboration in knowledge distillation.

Seghier, M.L., Kolanko, M.A., Leff, A.P., Jäger, H.R., Gregoire, S.M., Werring, D.J., 2011. Microbleed detection using automated segmentation (MIDAS): a new method applicable to standard clinical MR images. PloS one 6, e17547.

Shams, S., Martola, J., Cavallin, L., Granberg, T., Shams, M., Aspelin, P., Wahlund, L., Kristoffersen-Wiberg, M., 2015. SWI or T2*: which MRI sequence to use in the detection of cerebral microbleeds? the Karolinska Imaging Dementia Study. American Journal of Neuroradiology 36, 1089–1095.

Smith, S.M., 2002. Fast robust automated brain extraction. Human brain mapping 17, 143–155.

Sprigg, N., Flaherty, K., Appleton, J.P., Salman, R.A.S., Bereczki, D., Beridze, M., Christensen, H., Ciccone, A., Collins, R., Czlonkowska, A., et al., 2018. Tranexamic acid for hyperacute primary intracerebral haemorrhage (TICH-2): an international randomised, placebo-controlled, phase 3 superiority trial. The Lancet 391, 2107–2115.

Sundaresan, V., Arthofer, C., Zamboni, G., Dineen, R.A., Rothwell, P.M., Sotiropoulos, S.N., Auer, D.P., Tozer, D., Markus, H.S., Miller, K.L., et al., 2021. Automated detection of candidate subjects with cerebral microbleeds using machine learning. medRxiv.

Vadacchino, S., Mehta, R., Sepahvand, N.M., Nichyporuk, B., Clark, J.J., Arbel, T., 2021. Had-net: A hierarchical adversarial knowledge distillation network for improved enhanced tumour segmentation without post-contrast images. arXiv preprint 2103.16617.

Wang, C., Martins-Bach, A.B., Alfaro-Almagro, F., Douaud, G., Klein, J.C., Llera, A., Fiscone, C., Bowtell, R., Elliott, L.T., Smith, S.M., et al., 2021. Phenotypic and genetic associations of quantitative magnetic susceptibility in uk biobank brain imaging. bioRxiv.

Wang, S., Tang, C., Sun, J., Zhang, Y., 2019. Cerebral micro-bleeding detection based on densely connected neural network. Frontiers in neuroscience 13, 422.

Xie, Q., Luong, M.T., Hovy, E., Le, Q.V., 2020. Self-training with noisy student improves imagenet classification, in: Proceedings of the IEEE/CVF Conference on Computer Vision and Pattern Recognition, pp. 10687–10698.

Yang, C., Xie, L., Su, C., Yuille, A.L., 2019. Snapshot distillation: Teacher-student optimization in one generation, in: Proceedings of the IEEE/CVF Conference on Computer Vision and Pattern Recognition, pp. 2859–2868.

Yang, X., Zeng, Z., Yeo, S.Y., Tan, C., Tey, H.L., Su, Y., 2017. A novel multi-task deep learning model for skin lesion segmentation and classification. arXiv preprint 1703.01025.

Ye, J., Wang, X., Ji, Y., Ou, K., Song, M., 2019. Amalgamating filtered knowledge: Learning task-customized student from multi-task teachers. arXiv preprint 1905.11569.

Zhang, L., Song, J., Gao, A., Chen, J., Bao, C., Ma, K., 2019. Be your own teacher: Improve the performance of convolutional neural networks via self distillation, in: Proceedings of the IEEE/CVF International Conference on Computer Vision, pp. 3713–3722.

Zhang, Y., Brady, M., Smith, S., 2001. Segmentation of brain MR images through a hidden markov random field model and the expectation-maximization algorithm. IEEE transactions on medical imaging 20, 45–57.

Zhang, Y.D., Hou, X.X., Chen, Y., Chen, H., Yang, M., Yang, J., Wang, S.H., 2018. Voxelwise detection of cerebral microbleed in cadasil patients by leaky rectified linear unit and early stopping. Multimedia Tools and Applications 77, 21825–21845.

Zhang, Y.D., Hou, X.X., Lv, Y.D., Chen, H., Zhang, Y., Wang, S.H., 2016. Sparse autoencoder based deep neural network for voxelwise detection of cerebral microbleed, in: 2016 IEEE 22nd International Conference on Parallel and Distributed Systems (ICPADS), IEEE. pp. 1229–1232.

Zhou, G., Fan, Y., Cui, R., Bian, W., Zhu, X., Gai, K., 2018. Rocket launching: A universal and efficient framework for training well-performing light net, in: Proceedings of the AAAI Conference on Artificial Intelligence.

