## Supplementary Material for "Automated Detection of Cerebral Microbleeds on MR images using Knowledge Distillation Framework"

### 1 Comparison of existing patch-level CMB detection methods

Table S1: Comparison of the performance of the existing methods that were developed and evaluated at patch-level. Pl.TPR - patch-level TPR, Pl.Prec - patch-level precision, Pl.Acc - patch-level accuracy.

| **Methods** | **Sequence(s) (# test subjects)** | **Patch-size** | **# CMB/non-CMB patches** | **CMB patches detection performance** |
|---|---|---|---|---|
| Zhang et al., 2016 [1] (10-fold cross-validation) | SWI (5) | 20 × 20 | 30,478/30,711 | Pl.TPR - 93.2%, Pl.acc - 93.22% |
| Chen et al., 2018 [2] | SWI (12) | 16 × 16 × 8 | 377/1235 | Pl.TPR - 94%, FPavg - 11.7, Cl.Prec - 72% |
| Zhang et al., 2018 [3] (10-fold cross-validation) | SWI (20) | - | 68,847/68,829 | Pl.TPR - 93.05%, Pl.acc - 93.06% |
| Zhang et al., 2018 [4] (10-fold cross-validation) | SWI (20) | 61 × 61 | 68,847/68,829 | Pl.TPR - 95.13%, Pl.acc - 94.23% |
| Hong et al., 2019 [5] (10-fold cross-validation) | SWI (20) | 61 × 61 | 4287/4287 | Pl.TPR - 95.7%, Pl.acc - 97.4% |
| Wang et al., 2019 [6] (10-fold cross-validation) | SWI (20) | 61 × 61 | 10000/10000 | Pl.TPR - 97.8%, Pl.acc - 97.7% |
| Lu et al., 2021 [7] (5-fold cross-validation) | SWI (20) | 41 × 41 | 6407 / 6624 | Pl.TPR - 98.27%, Pl.acc - 98.60% |

### References


[1] Yu-Dong Zhang, Xiao-Xia Hou, Yi-Ding Lv, Hong Chen, Yin Zhang, and Shui-Hua Wang. Sparse autoencoder based deep neural network for voxelwise detection of cerebral microbleed. In *2016 IEEE 22nd International Conference on Parallel and Distributed Systems (ICPADS)*, pages 1229–1232. IEEE, 2016.

[2] Yicheng Chen, Javier E Villanueva-Meyer, Melanie A Morrison, and Janine M Lupo. Toward automatic detection of radiation-induced cerebral microbleeds using a 3d deep residual network. *Journal of digital imaging*, pages 1–7, 2018.

[3] Yu-Dong Zhang, Xiao-Xia Hou, Yi Chen, Hong Chen, Ming Yang, Jiquan Yang, and Shui-Hua Wang. Voxelwise detection of cerebral microbleed in cadasil patients by leaky rectified linear unit and early stopping. *Multimedia Tools and Applications*, 77(17):21825–21845, 2018.

[4] Yu-Dong Zhang, Yin Zhang, Xiao-Xia Hou, Hong Chen, and Shui-Hua Wang. Seven-layer deep neural network based on sparse autoencoder for voxelwise detection of cerebral microbleed. *Multimedia Tools and Applications*, 77(9):10521–10538, 2018.

[5] Jin Hong, Hong Cheng, Yu-Dong Zhang, and Jie Liu. Detecting cerebral microbleeds with transfer learning. *Machine Vision and Applications*, 30(7):1123–1133, 2019.

[6] Shuihua Wang, Chaosheng Tang, Junding Sun, and Yudong Zhang. Cerebral micro-bleeding detection based on densely connected neural network. *Frontiers in neuroscience*, 13:422, 2019.

[7] Si-Yuan Lu, Deepak Ranjan Nayak, Shui-Hua Wang, and Yu-Dong Zhang. A cerebral microbleed diagnosis method via featurenet and ensembled randomized neural networks. *Applied Soft Computing*, page 107567, 2021.
